# The impact of large mobile air purifiers on aerosol concentration in classrooms and the reduction of airborne transmission of SARS-CoV-2

**DOI:** 10.1101/2021.07.23.21261041

**Authors:** F. F. Duill, F. Schulz, A. Jain, L. Krieger, B. van Wachem, F. Beyrau

**Author notes:** <u>Corresponding author: F. F. Duill</u>, <u>Institut für Strömungstechnik und Thermodynamik</u>, <u>Otto-von-Guericke-Universität Magdeburg</u>, <u>Universitätsplatz 2</u>, <u>39106 Magdeburg</u>.

## Abstract

In the wake of the SARS-CoV-2 pandemic, an increased risk of infection by virus-containing aerosols indoors is assumed. Especially in schools, the duration of stay is long and the number of people in the rooms is large, increasing the risk of infection. This problem particularly affects schools without pre-installed ventilation systems that are equipped with filters and/or operate with fresh air. Here, the aerosol concentration is reduced by natural ventilation. In this context, we are investigating the effect of large mobile air purifiers (AP) with HEPA filters on particle concentration and their suitability for classroom use in a primary school in Germany. The three tested APs differ significantly in their air outlet characteristics. Measurements of the number of particles, the particle size distribution, and the CO_2_ concentration were carried out in the classroom with students (April/May 2021) and with an aerosol generator without students. In this regard, the use of APs leads to a substantial reduction in aerosol particles. At the same time, the three APs are found to have differences in their particle decay rate, noise level, and flow velocity. In addition to the measurements, the effect of various influencing parameters on the potential inhaled particle dose was investigated using a calculation model. The parameters considered include the duration of stay, particle concentration in exhaled air, respiratory flow rate, virus lifetime, ventilation interval, ventilation efficiency, AP volumetric flow, as well as room size. Based on the resulting effect diagrams, significant recommendations can be derived for reducing the risk of infection from virus-laden aerosols. Finally, the measurements were compared to computational fluid dynamics (CFD) modeling, as such tools can aid the optimal placement and configuration of APs and can be used to study the effect of the spread of aerosols from a source in the classroom.

## 1. Introduction

In the course of the SARS-CoV-2 pandemic, schools and other public institutions in Germany, as well as other countries, were closed because the risk of SARS-CoV-2 infection is considered significantly higher indoors than outdoors (Qian et al., 2021). The risk of infection also increases significantly with the residence time of the people in a room. According to (WHO, 2020) SARS-CoV-2 can be transmitted mainly in three ways. In addition to contact and droplet transmission as well as fomite transmission (Doremalen et al., 2020), airborne transmission is considered to play a crucial role. Some publications even assume a dominant role of virus-laden airborne particles for SARS-CoV-2 transmission in some cases (Ma et al., 2020; Morawska & Milton, 2020; Stadnytskyi et al., 2020; R. Zhang et al., 2020; Kriegel et al., 2020) and a subordinate role in others (Klompas et al., 2020; Kim et al., 2020).

Due to the large number of documented events in which infection with the Coronavirus occurred despite a large spatial distance between sender and recipient (Bae et al., 2020; Brlek et al., 2020; Cai et al., 2020; Charlotte, 2020; Groves et al., 2021; Hamner et al., 2020; Jang et al., 2020; Katelaris et al., 2021; Lendacki et al., 2021; Li et al., 2021; Lu et al., 2020; Mizumoto & Chowell, 2020; Moharir et al., 2021; Shen et al., 2020; Günther et al., 2020), as well as studies in which the viability of viruses in aerosols under laboratory conditions has been demonstrated to be as long as 16 hours (Fears et al., 2020) with a half-life of more than one hour (Doremalen et al., 2020), infection by viruses in airborne particles can be regarded as a serious route of spread. In their publication, signed by 239 scientists, (Morawska & Milton, 2020) urge more attention to be paid to the airborne spread of viruses.

Extensive flow simulations by the Technical University of Berlin, e.g. (HRI/TU Berlin, 2020), as well as by the RIKEN Center for Computational Science / Kobe University in Japan, e.g. (Makoto, 2020), show the distribution of exhaled particles and how they spread through the air (Kriegel & Hartmann, 2020). In addition to larger droplets that humans produce primarily when coughing and sneezing, exhaled particles during breathing and talking have a size of less than 5 µm. These particles can dehydrate to sizes of around 1 µm and less within a fraction of a second, depending on the temperature and humidity (Asadi et al., 2019). The exhaled air leads almost exclusively to particles smaller than 1 µm, a large part of which is even smaller than 0.3 µm (Fairchild & Stampfer, 1987; Papineni & Rosenthal, 1997). Particles of these sizes remain airborne for hours without sinking significantly. In a closed room with several people, this creates an atmosphere that enhances infection with SARS-CoV-2. The virus particle size itself is reported to be in the range of 0.08 - 0.14 µm (Laue et al., 2021). In rooms where multiple people are present for a long time, such as in schools, regular natural ventilation is a means of transporting potentially virus-contaminated aerosols out of the room or diluting them in such a way that the risk of infection is reduced. The German Federal Environment Agency, therefore, recommends ventilation phases that last 5 minutes and are to be initiated at 20-minute intervals (IRK, 2020). Depending on the aforementioned boundary conditions, fresh air can be supplied to the room to replace the potentially virus-laden room air.

However, regular cross-ventilation is associated with difficulties under real conditions at schools in the cold months, as the cooling of the room leads to thermal discomfort, which is exacerbated by wind speeds in the elevated range. A crucial practical problem of manual ventilation as an infection control measure is that it is hardly controllable and the correct and regular implementation cannot be ensured. In practice, natural window ventilation can therefore not be considered a safe measure for infection control without restrictions. Pre-installed ventilation systems or heating, ventilation and air conditioning (HVAC) systems could provide a remedy here, as they supply the room with conditioned, preheated fresh air. Yet, such ventilation systems are only installed in about one in ten schools in Germany (VDMA, 2020) and installation for every classroom is certainly not feasible in the short term.

Another way to reduce aerosol concentration indoors is the use of mobile air purifiers (AP). These APs work in recirculation mode. The room air is sucked in and passed through filters in which aerosol particles are separated from the air before the cleaned air is returned to the room. If filters of the HEPA class are used in APs, for example, 99.95 % (H13) of the aerosol particles can be removed. There are also APs that work with UV-C radiation or ozone, plasma, or ionization. In this study, we focus only on devices with HEPA filters.

The difference between APs and pre-installed ventilation systems or HVAC systems is that the APs do not supply fresh air to the room. The air humidity and the CO_2_ concentration in the room are therefore not changed. This means that APs can only be seen as an additional measure alongside ventilation and cannot be used as a stand-alone, independent measure. The use of APs as an additional measure is called for by researchers from the German Society for Aerosol Research (GAef e.V.), among others (GAeF, 2020)(GAeF, 2021). Until the beginning of July 2021, the German Federal Environment Agency considered the purchase of such air purifiers for schools to be useful only in exceptional cases and referred to insufficient studies and test evidence on the effectiveness of APs under realistic conditions (IRK, 2020). Furthermore, a guideline of the leading German Societies for Epidemiology (DGEpi), for Public Health (DGPH), for Paediatrics and Adolescent Medicine (DGKJ), and for Paediatric Infectiology (DGPI) points out the proportionality of acquisition costs and maintenance effort to the presumed positive effect, which results in the assessment that “neither the positive nor the negative effects” predominate (Leitliniengruppe Deutschland, 2021). According to an updated statement of 9^th^ of July 2021, professional APs for schools are now considered sensible by the German Federal Environment Agency “in order to minimize the probability of indirect infections during the pandemic” (UBA, 2021).

There are only a few experimental studies on the effectiveness of APs with HEPA filters under real conditions. (Küpper et al., 2019) have investigated portable APs in office spaces. In a music classroom, portable APs were investigated by (Narayanan & Yang, 2021). Measurements in school classrooms were also carried out by (Polidori et al., 2013) who investigated the positive influence on the reduction of particulate matter, as well as (Park et al., 2020). (Jhun et al., 2017) were also able to demonstrate a significant reduction in particulate matter and black carbon (BC) in classrooms; the researchers particularly focus on the positive effects for asthmatics.

In the wake of the COVID 19 pandemic, APs gained increased attention because they can be retrofitted without structural changes, making them easy to install and eliminating the need for costly installation as with pre-installed ventilation systems or HVAC systems. A study conducted by (Curtius et al., 2020) in a school in Frankfurt a.M. in Germany shows the positive influence of APs on the reduction of aerosol concentration in indoor air. In the study, four small mobile APs were used, which were able to significantly reduce the aerosol concentration in a classroom. Thus, the aerosol concentration at the measuring points could be reduced by 90 % in less than 30 minutes. The measurements by Curtius and his team cover the particle size classes < 3 nm to 10 µm. Furthermore, the particulate matter PM_10_ and the CO_2_ concentration were measured during school operation. Based on a computational model by (Lelieveld et al., 2020) and the measurements of the study, it was determined that the potential amount of virus-laden particles inhaled by people in the room is reduced by a factor of 6 through the use of APs. Here, a highly infectious person was assumed to spend two hours with other people in a room where APs with a total air-circulation rate of 5.7 h^-1^ were in operation. In a study by (Burgmann & Janoske, 2021) a large floor-standing AP was investigated in a classroom, which was operated on the short side of the room with a volume flow of 1200 m^3^/h. The aerosol concentration and distribution were determined in order to validate simulations that had also been carried out.

In addition to the small mobile APs used in the study by Curtius and his team, towards the end of 2020 the first large stand-alone units were also available as in the study by (Burgmann & Janoske, 2021), of which only one unit is needed to achieve air circulation rates of 5 - 6 h^-1^ in standard classrooms (50 – 80 m^2^). The large APs have lower noise levels than the small units (e.g. the units used in (Curtius et al., 2020)). At the same time, several small units must be used to achieve air circulation rates of 5 – 6 h^-1^, which in turn leads to an increase in the noise level.

The large floor-standing units are characterized by the fact that the discharged air flows out horizontally at a height of more than 2 meters or is directed towards the ceiling at a lower discharge height. This generates a flow along the ceiling, which leads to large-area distribution of the filtered air, reducing the aerosol concentration in the room as homogeneously as possible. The air intake is at floor level. Such a floor-standing unit, which has the outlet of the filtered air mainly to the sides and is placed centrally on the wall opposite the blackboard on the short side, was investigated by (Burgmann & Janoske, 2021).

In the present study, three different types of the described large floor-standing APs are to be tested in real school operation in a primary school. The main difference between the three floor-standing units used is the direction in which the purified air is discharged. The aim is to determine whether even a single AP can homogeneously and significantly reduce the aerosol concentration in the entire room without negatively influencing the teaching process, e.g. through a high noise level. Furthermore, the decay rate of the APs is examined and classified under the same, reproducible conditions using an aerosol generator. For one of the devices, the room position was varied, as well. A simplified calculation model is applied to assess the influence of various parameters such as room size, duration of stay, and ventilation efficiency. With the help of the resulting effect diagrams, it is possible to compare conventional window ventilation with the AP application. At the same time, the effect diagrams allow us to draw conclusions about the impact of various influencing parameters on the risk of infection. Finally, the optimal placement and operating conditions of the APs are researched using computational fluid dynamics (CFD) tools.

## 2. Setup

This section provides a detailed description of the investigated indoor APs and the measuring instruments used. In addition, the classrooms and the location of the APs are specified and the scenarios that were investigated within the scope of this study are defined.

### 2.1 Mobile air purifiers used/requirements for air purifiers for schools

Three different types of mobile APs were used for the study, which are large floor-standing units (floor height > 1.8 m) equipped with HEPA filters. All three AP types have an air intake at floor level. In all devices, the room air is first passed through a pre-filter, whereby the corresponding filter area varies in size between the units (see Table A 2). This pre-filter is intended to separate coarser particles and thus protect the main filter against rapid clogging. The main filters, which are installed behind the pre-filters, are HEPA filters that have an efficiency of at least 99.95 % for H13 (or 99.995 % for H14) at 0.1 - 0.3 µm (MPPS). The areas of the filters, as well as data on the filters, are summarized in Table A 2 in the Appendix. According to VDI standard 6022 (VDI 6022, 2018), the pre-filters must be replaced annually and the HEPA filters every two years. All three AP types operate with an EC fan.

The three devices differ mainly in their air outlet. In all cases, this takes place at a height above 1.7 m. AP 1 is equipped with three air ejection nozzles, which are mounted on the front at a height of approximately 1.7 m and whose ejection directions are individually adjustable (see Figure A 1 a.). AP 2 and 3 have the air outlet at a height of about 2.3 m. In both cases, the output of the filtered air is directed horizontally with a slight upward angle. In AP 2, the air is mainly discharged to the sides. The air intake at floor level is also on the sides (AP 2), whereas AP 1 and AP 3 take in air from all sides. AP 3 has an air outlet that is directed equally to the front and the sides. These different air outlet variants result in different optimal locations in the room. For AP 1, this is centrally located on the short side of the room opposite the blackboard, as the forward-directed flow can generate a room roll over the long side of the room. AP 2, on the other hand, should be placed centrally on the long side of the room due to the lateral air outlets, as the manufacturer also recommends. AP 3 can be positioned centrally on both the short and the long sides of the room wall. This applies to the installation of one unit per room. Due to the high-volume flows (> 1000 m^3^/h) that the units can provide, one unit per room is sufficient for the classroom size (186 m^3^) of this study.

The tested devices achieve volume flows of over 1000 m^3^/h (see Table A 2 in the appendix). The volume of the room and the number of people in it are decisive for determining the volume flow to be set for the respective classroom. Concerning the room volume, an air circulation rate of 5 - 6 h^-1^ should be aimed for (IRK, 2020), whereas the WHO refers to DIN EN 16798-1 and recommends a minimum air volume of 36 m^3^/h per person (WHO, 2021). Especially in large rooms, a person-related circulation rate is preferable to a volume-related one, because in large rooms a stronger dilution of the aerosol particles can take place. The circulation rate is therefore not always relevant (Kriegel, 2020).

For the comparative studies, the units are operated in teaching mode at 1000 m^3^/h (AP 2, AP 3) and 1060 m^3^/h (AP 1). This corresponds to an air circulation rate of 5.4 h^-1^. For use in ongoing teaching mode, the noise level must be given special attention. According to the German Federal Institute for Occupational Safety and Health “Arbeitswissenschaftliche Erkenntnisse” AWE 124 (Probst, 2003), a sound pressure level of 35 – 40 dB(A) should not be exceeded “in the case of very high concentration requirements, such as for demanding material processing, programming or scientific work”. VDI Guideline 2081 (VDI2081, 2019) sets a maximum sound pressure level of 35 dB(A) for classrooms, as does the German Workplace Guideline (ASR A3.7, 2018). The Commission on Indoor Air Hygiene (IRK) at the Federal Environment Agency in Germany considers noise levels above 40 dB(A) to be disruptive to teaching (IRK, 2020).

The sound level of the units was calculated under various boundary conditions by the manufacturers or by institutes commissioned to do so. For APs 1 and 2, the sound levels at a distance of 1 m were calculated with the influence of the room for an average reverberation time of 0.5 s. In the case of AP 3, 8 dB(A) was subtracted from the sound power, which corresponds to the sound level in the free field at 1 m distance. The values calculated in this way can be found in Table A 2 of the appendix. In order to be able to compare the sound pressure levels under real conditions, measurements were taken on all three units under the same conditions as part of this study, which can be found in chapter 3.5.

### 2.2 Measurement equipment

#### Aerosol spectrometer

Two aerosol spectrometers of the type AQ Guard from PALAS GmbH were used to measure the air quality. The measuring device is designed for indoor use and analyses airborne particles in the size range 0.178 - 17.78 µm in 64 size classes in continuous operation. Particle measurements in the optical aerosol spectrometer are carried out by means of scattered light analysis on individual particles. For this purpose, the individual particles are passed through a defined measuring volume, which is homogeneously illuminated by a polychromatic LED light source. The number of particles per time unit is then determined on the basis of the scattered light pulses generated by the particles. The measuring range of the particle number concentration lies between 0 - 20,000 particles/cm^3^. In terms of mass, the measuring range is specified by the manufacturer as 0 - 20,000 µg/m^3^.

In addition to the number of particles and the particle size distribution, the fine dust classes PM_1_, PM_2.5_, PM_4_, PM_10_, PM_tot_ (TSP) as well as pressure, temperature, relative humidity, CO_2_, and volatile organic hydrocarbons (TVOC) are recorded. The CO_2_ concentration measurement takes place in the range of 0 – 5000 ppm via an NDIR sensor, with an accuracy of ± 30 ppm. A list of the instrument’s specifications can be found in Table A 1.

#### Aerosol generator

The PAG 1000 from PALAS GmbH was used to generate an aerosol. This is a portable aerosol generator that can provide a volume flow of 0.9 - 4.6 l/min. The device can work with di-ethyl hexyl sebacate (DEHS) and similar oils as well as NaCl and KCl. DEHS was used in this study. The flow rate of the aerosol generator can be adjusted to two levels (Low, High), each from 0 - 100 %. For example, when using DEHS, at the high level and 100 % (4.6 l/min) 1.2*10^9^ particles/s (1.6*10^7^ particles/cm^3^) with a size of ≥ 0.2 µm are generated.

#### Sound analyzer

In order to be able to measure the noise level of the units, a 2260 Investigator module sound analyzer from Brüel & Kjær was used with the BZ7201 sound analysis software. This is a class 1 sound level meter (IEC and ANSI). The microphone used was a Brüel & Kjær type 4189 permanently polarised free-field microphone. The equivalent continuous sound level was measured for 20 seconds.

### 2.3 Set-up of the test series

#### Location of the air purifier

If a large floor-standing device is chosen, as in this study, the location in the room is crucial for the effectiveness of the unit. For the large professional units, a location on the wall should be chosen. On the one hand, this is important for safety reasons, as securing to the wall is necessary in schools for devices that are larger than the students themselves. On the other hand, the air outlet on the APs is also designed for positioning on the wall. This refers to normal classroom sizes, where a unit can generate an air circulation rate of 5 - 6 h^-1^.

The location of the AP in a classroom is further dependent on various factors:

- Room geometry
- Air outlet direction of the AP
- Conditions of the room ceiling (e.g. protruding lamps, crossbars)

Especially with APs 2 and 3, where an air outlet is to the side, a central location on one of the wall sides is advantageous from a fluidic point of view. Positioning in a corner should be avoided. However, if the APs are integrated into a classroom, modifications to the existing furniture may be necessary.

Figure 1 shows the locations and the corresponding outlet of the filtered air. The flow spreads shown there are intended to give a qualitative impression of the discharge direction. No direct statement about the throw or the room flow can be derived from this. The qualitative characterization of the flow on which the illustration is based was carried out using a fog generator.

**Figure 1:**
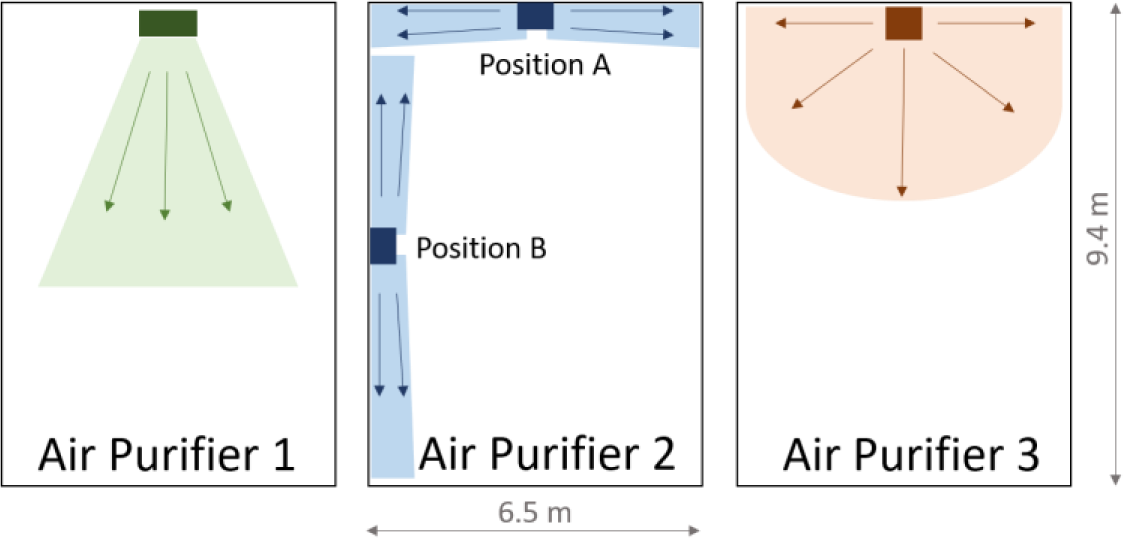
Illustration of the installation position of the three different APs used including the qualitative flow at the outlet. AP 2 was examined individually at two different locations (A and B).

#### The layout of the classroom

The APs investigated in this study are located in different classrooms of a primary school. The dimensions of the classrooms in which the tests took place are identical at 9.4 x 6.5 m and are shown in Figure 2. The ceiling height is 3.05 m. The area of the classrooms is therefore 61.1 m^2^ and the volume 186.4 m^3^. The number, size, and orientation of the windows are also identical in all classrooms. The side of the room with the windows consists of four window areas, which in all classrooms face a side street with little traffic. Each of the window areas has two double casement windows arranged one above the other. During cross-ventilation in class, the bottom right window of the double casement window is fully opened. The area for window ventilation is therefore 2.324 m^2^. During window ventilation, the door was always opened as well as the windows in the adjacent corridor.

**Figure 2:**
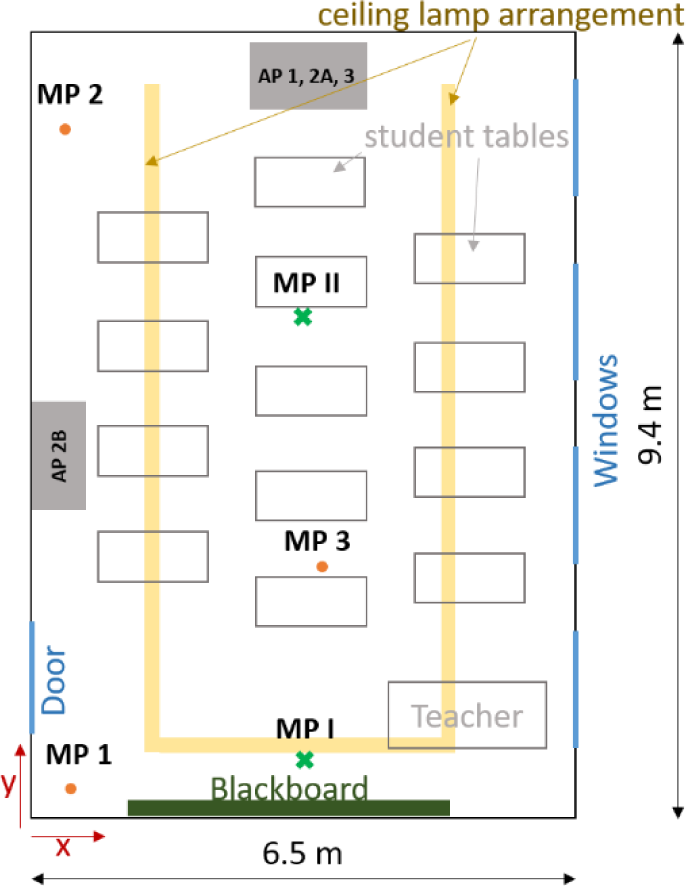
Sketch of a classroom with tables and locations of the APs including the positions of 5 different measuring points (MP 1, MP 2, MP 3, MP I, MP II).

In the classrooms, there are cupboards, desks, and ceiling lights. These are arranged almost identically in the different classrooms. Only the individual student desks have marginally different locations in the different classrooms, with all students in the sitting position facing the blackboard/classroom front. The rows of lamps have an angular ‘U’ arrangement, as shown in Figure 2. The open side is the short, backside of the room. The row of lamps is not flush with the ceiling but is suspended by 20 cm. This lamp arrangement favors the positioning of the APs on the rear, the short side of the room, as the airflow can thus exit into the room without obstruction.

### 2.4 Definition of the scenarios

Within the framework of the study, two different test scenarios were investigated. In scenario 1, measurements were carried out with the aerosol spectrometer during ongoing teaching. In scenario 2, a DEHS aerosol was fed into the deserted room by means of an aerosol generator, which makes it possible to obtain statements regarding the separation effect of particles of the individual devices tested.

#### Scenario 1

The measurements in the class took place in different rooms of a primary school with the same geometry. Measurements were taken with all three APs. AP 2 was tested at two different positions (Figure 2). A school lesson has a duration of 80 minutes. During the lesson, 5 – 22 students, a teacher, and, in some measurements, a research assistant who controlled the window ventilation were present. The measurements took place in April and May 2021.

In all measurements, which were performed with a scientist on site, ventilation was provided every 20 minutes for 5 minutes. During window ventilation, the door located on the other side of the room was also opened. The windows in the hallway were also open. For the measurements with cross-ventilation, a distinction is made between scenario 1.1 - with AP - and scenario 1.2 - without AP. In the case of classroom measurements without scientists, teachers took over the window ventilation, which varied depending on the weather and the teacher, but thus reflected real conditions. The measurements that took place without ventilation controlled by scientific staff are referred to in the following as scenario 1.3.

#### Scenario 2

In order to be able to determine the decay behavior of the aerosol concentration due to the use of the APs, measurements were carried out without students. For the defined addition of aerosol particles, an aerosol generator (see section 2.2 Measurement equipment) was used here, which was operated for 15 minutes with a volume flow of 4.6 l/min. While the generator was running, the aerosol particles were homogeneously distributed in the room by a fan. After 15 minutes, the generator was switched off and the particle concentration was homogenized in the room by means of a fan for another three minutes. After that, the AP was turned on and the measurement started by means of aerosol spectrometers at two different measuring points (MP). The aerosol spectrometers were positioned at the rear of the room at a distance of approximately 3 (MP 2) and 8 meters (MP 1) from the AP at a height of 0.95 m (Table 1, Figure 2). The measurement duration was 40 minutes in each case. These tests were carried out with all three APs, with AP 2 in the rear and side position (position A and B). In order to detect a potential influence of convection currents in the room as well as the deposition on surfaces on the measurement results, measurements were also carried out without APs. During these 40-minute measurements, no person was present in the room and the windows and door were closed. Each measurement was repeated three times. A volume flow of 1000 m^3^/h was set for AP 2 and 3, and 1060 m^3^/h for AP 1. The volume flow rate information comes from the manufacturers and was not checked during the investigation.

**Table 1:** Coordinates of the measuring points where z represents the height.

|  | x in (m) | y in (m) | z in (m) |
| --- | --- | --- | --- |
| **Scenario 1:** |  |  |  |
| MP1 | 0.99 | 0.18 | 1.73 |
| MP2 | 0.30 | 8.85 | 0.85 |
| MP3 | 3.50 | 4.80 | 0.92 |
| **Scenario 2:** |  |  |  |
| MP I | 3.25 | 0.70 | 0.95 |
| MP II | 3.25 | 6.00 | 0.95 |

In the context of scenario 2, sound level measurements were also taken at a distance of 2.5 m frontally in front of the respective APs at a height of 1.1 m (approximate head height of a seated student). One measurement took place over 20 seconds and forms the mean value of this period. Furthermore, the measurements per unit type (1, 2, 3) were carried out on two different units of each type to exclude possible production-related fluctuations. These measurements were also repeated three times each. The measurements in scenario 2 thus allow statements to be made about the suitability of the units for use in classrooms.

## 3. Results and Discussion of the experimental investigation

The measurement results shown and discussed in this section include the aerosol concentration in real school operation (scenarios 1.1, 1.2, and 1.3) as well as the corresponding CO_2_ concentrations. Scenario 1 includes measurements with (1.1 and 1.3) and without (1.2) APs as well as with (1.1 and 1.2) and without (1.3) window ventilation. The particle size distribution is measured in the range of 0.178 - 17.78 µm. In the measurement series without students (scenario 2), where an aerosol generator was used as a particle source, the decay rates of the tested APs are evaluated and compared with the case without AP. Furthermore, the sound levels of the three devices are measured.

### 3.1 Aerosol concentration in school operation - Scenario 1

The results discussed below are based on scenarios 1.1, 1.2, and 1.3 described in section 2.4, in which the aerosol concentration was measured in classrooms during ongoing teaching. The measurements in scenarios 1.1 and 1.2 were carried out with and without AP with simultaneous regular ventilation. The measurements in scenario 1.3, on the other hand, were carried out with AP but without ventilation. The upper diagram in Figure 4 shows a characteristic measurement curve of the aerosol number concentrations in the range 0.178 - 17.78 µm at a measurement point over a school lesson (duration 80 min) as well as the 15 minutes before and after the start of the lesson. The measurement curves of the aerosol number concentration of the three cases described (1.1, 1.2, 1.3) are shown in the upper diagram of Figure 4. AP 1 was used for the measurements shown in the figure.

The people in the room produce respiratory particles by breathing and speaking, and at the same time, they inhale particles. The amount of inhaled and exhaled particles does not differ significantly. Thus, the respiratory particles are not measurable by humans in this scenario and therefore they cannot be identified in the upper diagram of Figure 4. What is measurable, however, is the particle input of the people entering the class, which is depicted in the first 15 minutes of the diagram. The particle input can be identified with (blue curve) and without AP (red curve) by an increase in the number of particles; only in the case of the black curve, where the AP was also switched on, the curve stagnates. This is due to the fact that the students in the three scenarios shown did not enter the room in a comparable manner; in the case of scenario 1.3, students were already in the classroom 15 minutes before the start. Between the start of the lesson (minute 15) and the first window ventilation at minute 35, a reduction in the number of particles can be seen in the black and blue curves, which represent the scenarios with APs. Even in scenario 1.2, where no AP was used, a slight decrease in the number of particles can be detected, which is mainly due to deposition on surfaces. Sedimentation and coagulation processes also play a role as well as inhalation by people. Window ventilation, which was carried out in cases 1.1 and 1.2, leads to a significant particle input, which is noticeable at minute 35 by a slope (red and blue curve). The particle sizes introduced into the room are discussed in section 3.4. At minute 40, the window is closed. This leads to an immediate significant decrease in the measured particle number, in the measurement with and without the AP, which is again due to the processes mentioned above. The phase of rapidly decreasing particle number lasts 3 - 4 minutes, which in the case without AP is followed by a phase in which the particle number decrease is more moderate until it stagnates (50 - 60 min). In case 1.1 (blue curve), on the other hand, a strong decrease in the number of particles can still be seen, which continues until the next ventilation phase at minute 60 and can be attributed to the AP. The subsequent five-minute window ventilation restarts the process described above and does not differ significantly in its effect on the aerosol number concentration. The black curve differs from cases 1.1 and 1.2, which are significantly characterized by window ventilation. Since no window ventilation was performed in scenario 1.3, a continuous drop in the particle number concentration can be observed after the start of the lesson at minute 15, which flattens out with increasing time and approaching the x-axis. From minute 60 onwards, an approximately stagnating particle number concentration of fewer than 5 particles/cm^3^ is observed. However, this scenario must be viewed critically, as the CO_2_ concentration and air humidity rise continuously and the air quality is, therefore, unsuitable for teaching (see section 3.3).

No significant difference in the particle number curves can be detected for the different measuring positions. The measured values shown in the upper diagram of Figure 4 are taken from measuring point 1 (Figure 2) and thus represent the measuring point that is located furthest away from the AP.

In summary, a reduction of the particle number can be assumed due to the AP. Without the supply of particles through window ventilation, a low particle count in the single-digit range (particles/cm^3^) can be maintained. The significance is limited, however, as it is not possible to distinguish between respiratory particles and particles that are brought in with the fresh air.

### 3.2 Aerosol concentration measurements with aerosol generator, without students

In order to be able to investigate the influence of the investigated APs on the aerosol concentration in a definite and reproducible way without external influences, a DEHS aerosol was introduced into the room empty of people in scenario 2. The particle number concentration in the room was thus set approximately homogeneously to 6000 particles/cm^3^. Two aerosol spectrometers recorded the measured values. The diagrams in Figure 3 show these measurement results with the respective decay curves for measurement points I and II (see Figure 2); the values with the respective range from three measurements in each case can be found in Table 1.

**Figure 3:**
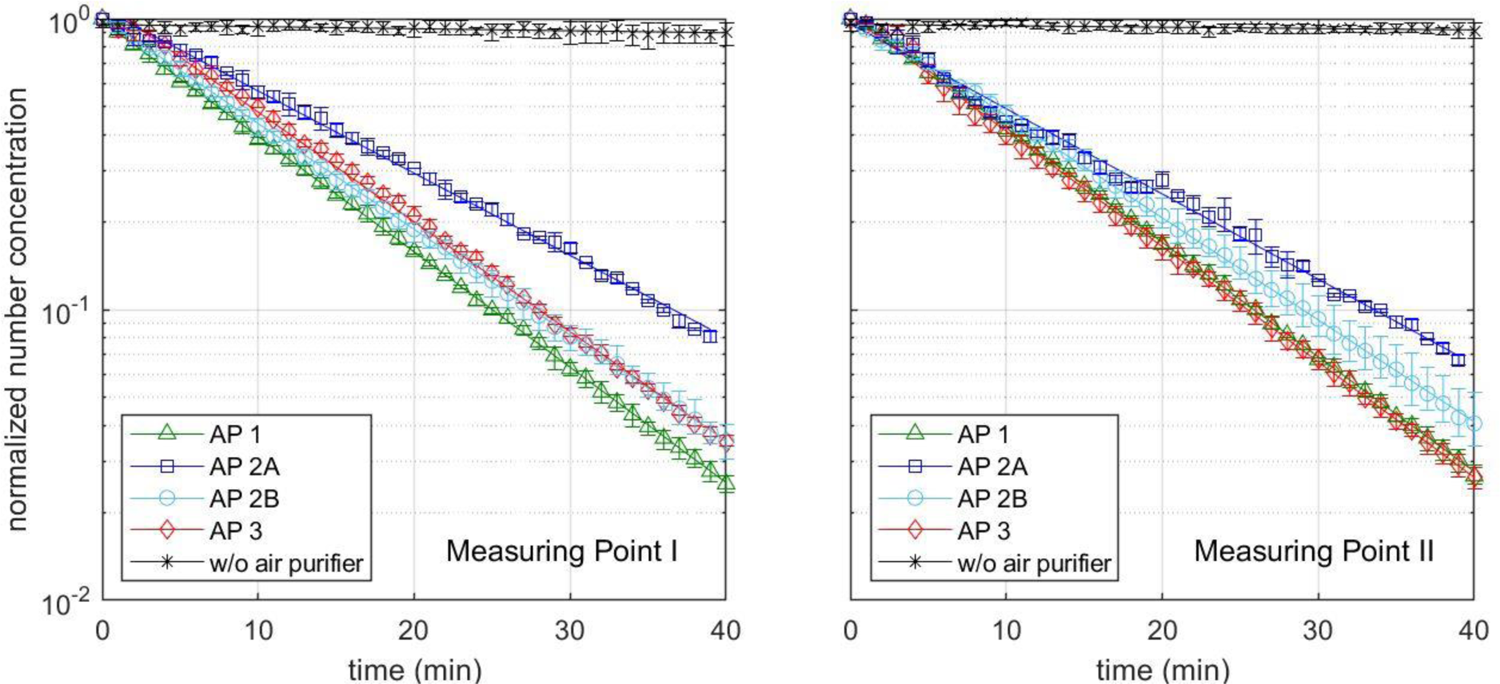
Normalized aerosol number concentration of three different APs at an air exchange rate of ∼5.4, as well as without AP at two different measuring points (MP I and MP II). AP 2 has been tested at two different positions, on the short side of the room (A) and on the long side of the room (B).

**Figure 4:**
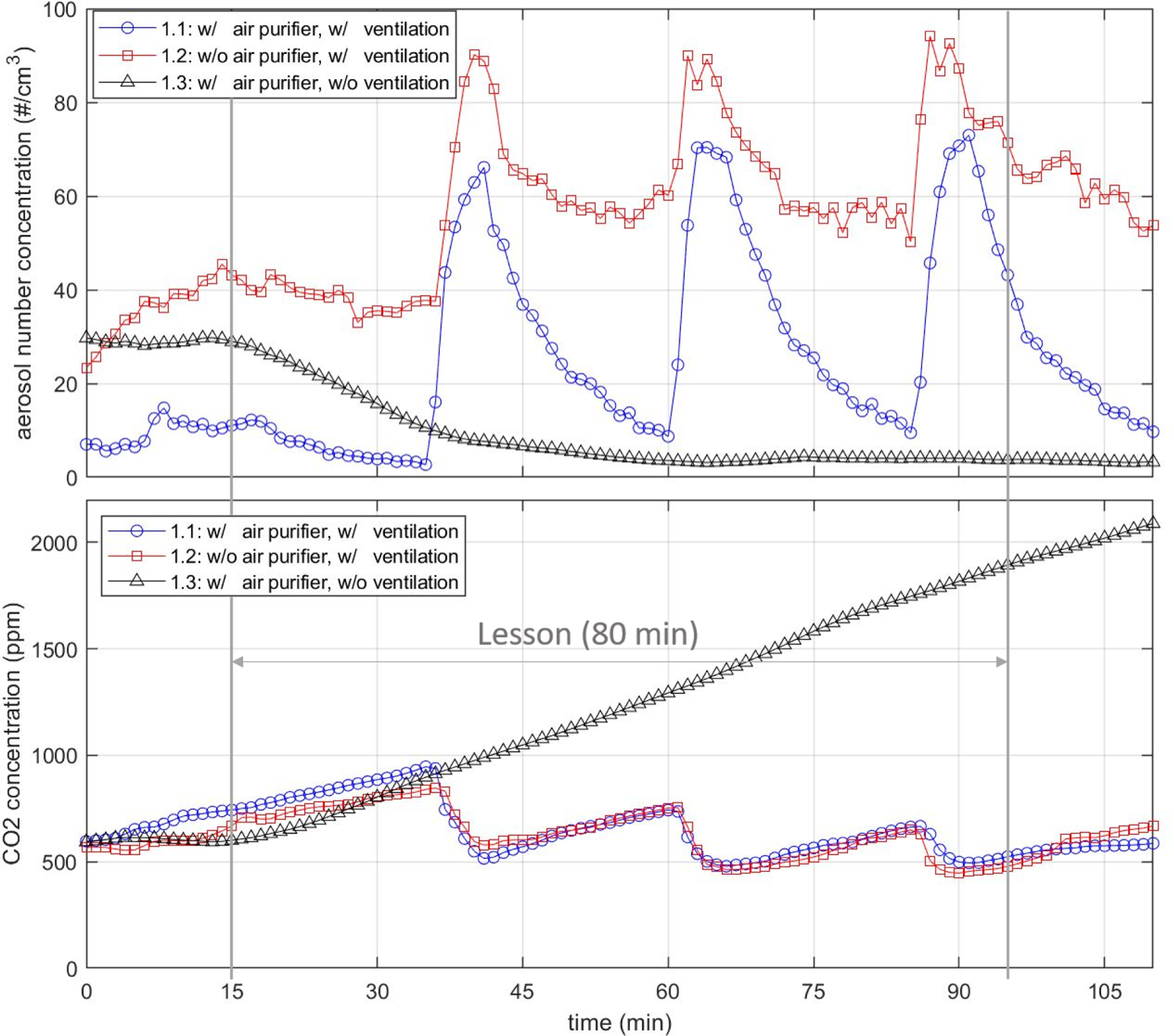
Aerosol number concentration (top) and CO_2_ (bottom) concentration for three different scenarios. The first scenario (blue) includes an AP at an air exchange rate of 5.4 h^-1^ and with manual ventilation every 20 minutes for 5 minutes. The second scenario (red) keeps the ventilation but with AP switched off. In the third scenario, the AP works again at an air exchange rate of 5.4 h^-1^, but without manual ventilation. The lesson started at minute 15 and ended at minute 95

The decay rate *λ* describes the negative slope of the logarithmic decay curve (lines) shown in Figure 3 according to the following decay function:

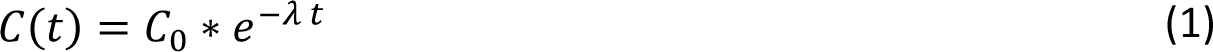

*C* represents the aerosol number concentration at the beginning (0) and at time *t*. The markers in Figure 3 show the mean values, whereby these are provided with error bars of the maximum and minimum measured values. The half-lives and the time after which 90 % of the particles were removed by the filters were determined in the following using the decay rate according to equation 1 (Table 2).

**Table 2:** Values of the decay rates, half-life, and time after 90 % of the aerosols have been removed for different APs at a set volume flow of ∼ 1000 m^3^/h and without AP considering two different measuring points as shown in Figure 2.

| | Set volume flow rate | Measuring Point | Decay rate ($\text{h}^{-1}$) | Range ($\text{h}^{-1}$) | Half-life (min) | 90 % removed in (min) |
| --- | --- | --- | --- | --- | --- | --- |
| AP 1 | 1060 $\text{m}^3/\text{h}$ | MP I | 5.48 | 0.20 | 7.59 | 25.21 |
|  |  | MP II | 5.44 | 0.26 | 7.65 | 25.40 |
| AP 2A | 1000 $\text{m}^3/\text{h}$ | MP I | 3.91 | 0.05 | 10.64 | 35.33 |
|  |  | MP II | 4.05 | 0.04 | 10.27 | 34.11 |
| AP 2B | 1000 $\text{m}^3/\text{h}$ | MP I | 5.03 | 0.38 | 8.27 | 27.47 |
|  |  | MP II | 4.85 | 0.61 | 8.58 | 28.49 |
| AP 3 | 1000 $\text{m}^3/\text{h}$ | MP I | 5.21 | 0.08 | 7.98 | 26.52 |
|  |  | MP II | 5.48 | 0.09 | 7.59 | 25.21 |
| w/o air purifier | - | MP I | 0.10 | 0.28 | 415.89 | 1381.55 |
|  |  | MP II | 0.07 | 0.11 | 594.13 | 1973.64 |

Without an AP, the particle number concentration decreases only marginally over the measuring period of 40 minutes at both measuring points (see Figure 3) with a decay rate in the range of 0.1 h^-1^. The reduction of particles can be attributed to coagulation processes and deposits on surfaces. The indication of a half-life and the time after 90 % of the particles have been removed cannot be obtained from the measurement results. With the decay rate, values of more than 400 minutes (half-life) or more than 1350 min (reduction by 90 %) can be calculated (see Table 2). If an AP is used at 1060 (AP 1) or 1000 m^3^/h (AP 2, AP 3), the decay rates increase considerably. Among the devices used in this study, AP 2 at position A achieves the lowest decay rates of 3.91 h^-1^ (MP I) and 4.05 h^-1^ (MP II) with only a slight difference between the measuring points. For the half-lives, this means values of 10 – 11 minutes (Table 2), for the removal of 90 % of the particles present at the beginning 34 – 35 minutes. It should be mentioned that the air inlet, as well as the air outlet, is on the side. At a distance of 0.3 m from the air inlet, there is furniture on both sides (see Figure A 1 b.). The furniture is also next to AP 1 and AP 3, however, both devices have a 360° intake. If this furniture is removed, the decay rate of AP 2 at position A increases by 15 % at the measuring point on the other side of the room (MP I) (see Figure 10) and 2 % at the measuring point near the unit (MP II). If the device is positioned on the long side of the room opposite the windows (position B), the decay rate increases to 5.03 h^-1^ (MP I) and 4.85 h^-1^ (MP II). The half-lives and the time for the separation of 90 % of the particles are reduced by almost 20 % compared to location A (Table 2). AP 1 and 2 have an almost identical decay rate of 5.44 h^-1^ (AP 1) and 5.48 h^-1^ (AP 3) at measuring point II, which is closer to the AP, so that the half-lives for both are less than 8 minutes (Table 2) and 10 % of the initial concentration is reached after less than 26 minutes in each case (Table 2). At measuring point I on the opposite side of the room to the AP, the decay rate of AP 1 with 5.48 h^-1^ is slightly higher than that of AP 3 with 5.21 h^-1^, which may be due to the nozzles of AP 1, which are directed exclusively to the front and achieve a targeted dilution of the air to the front. The nozzle arrangement of the AP 1 generates a greater throwing distance due to the focused airflow. Thus, AP 1 achieves a half-life of 7.59 min, the AP 3 7.98 min. 90 % of the particles were removed after 25.4 (AP 1) and 26.52 min (AP 3). However, a direct comparison based on the decay rates of the different units is only possible to a limited extent, as the actual volumetric flows of the units were not measured before the tests. Nevertheless, together with noise level measurement, the decay rate can have a high informative value for the suitability of APs in classrooms.

In summary, it can be concluded that the results of the measurements with the different APs show small fluctuation ranges in each case, and the reproducibility is assessed as good. Contrary to the possible expectation that the effect of an AP depends only on its volume flow, the results show that the use of APs with almost the same volume flow leads to different results of air purification. In the present case, this is mainly due to the resulting room flow with the associated recirculation zones. The furniture next to the equipment can also show an influence on the results (see Figure 10). A beneficial design of the air outlets and the air intake, as in AP1 and AP3, have a significant influence on the results here.

### 3.3 CO_2_-Concentration

The CO_2_ concentration in classrooms is an important parameter, with the help of which statements can be derived on the spread and distribution of breathing air in rooms. CO_2_ measurements thus allow conclusions to be drawn about the course of the potential virus concentration in rooms without the operation of an AP and CO_2_ concentration makes it possible to record the influence of window ventilation on air exchange. In addition, CO_2_ has a physiological effect that requires special attention in connection with the learning environment of children.

In Germany and throughout Europe, there are no binding regulations for maximum permissible CO_2_ levels in public buildings. Guidelines, such as the Workplace Directive 3.6 in Germany, define good indoor air quality as < 1000 ppm (Pettenkofer, 1858). Exceeding 1500 ppm can lead to a reduction in general well-being, drowsiness, a decrease in the ability to concentrate, or headaches, depending on the person and their state of mind. The average CO_2_ concentration in the atmosphere is 419 ppm (as of April 2021).

APs do not influence the CO_2_ concentration or the air humidity, so even when using APs, regular ventilation is necessary to improve the air quality. The influence of window ventilation can be seen in the CO_2_ measurement data. The lower diagram in Figure 4 shows two typical CO_2_ curves for cases 1.1 (blue) and 1.2 (red). In this case, controlled window ventilation was initiated for 5 minutes after every 20 minutes during the lesson. In the diagram, it should be noted that a period of 15 minutes was already recorded before the start of the 80-minute lesson. The actual lessons take place in the period from 15 to 95 minutes. In the red and blue graphs, the ventilation intervals were identical in each case; in the blue curve, an AP was in operation at the same time. The third, black graph (case 1.3) shows the CO_2_ concentration for a lesson in which no window ventilation took place. Here an almost linear CO_2_ increase can be seen from the beginning (15 min) to the end (95 min) of the lesson. The increase depends on the number of people in the room and their activity. In case 1.3 (black) there were 22 people, in case 1.1 (blue) and 1.2 (red) 7 people in the classroom. The increase in case 1.3 is therefore comparatively steeper.

The measurement results of scenarios 1.1 and 1.2 show that the CO_2_ concentration can be kept below 1000 ppm with regular cross-ventilation. It must be taken into account, that the number of pupils present during the measurements was only one-third of the usual number due to the pandemic-related alternating lessons. Scenario 1.3, on the other hand, shows another practical case that can occur more often during the cold season. This assertion is substantiated by measurement results from this study and also supported by field reports from the authors’ environment. During this measurement campaign, outside temperatures were around 0°C and there was a strong wind at the same time. No academic staff member was in the classroom during the measurements. The window ventilation was therefore controlled by the teacher, just as in real schools. According to the measurement data, no window ventilation took place during the school stand in these cold weather conditions. In this context, it should be noted that ventilation at low outdoor temperatures usually has a negative impact on thermal comfort. In cases where ventilation is insufficient, the operation of an AP is particularly effective in reducing the number of particles. This is because, without sufficient air exchange through manual ventilation, the virus concentration introduced into the room by an infectious person would continuously increase without the operation of an AP, which would also increase the risk of infection for persons present. The effect is illustrated by the black graph of particle concentration in Figure 4. Here, the aerosol concentration remains at a constantly low level, even if the air quality is hygienically questionable due to high CO_2_ values and high humidity.

### 3.4 Particle size distribution

In the course of the measurements in class, which extended over 10 school days, very small particle sizes, in the measuring range of 0.178 - 17.78 µm, dominated in terms of their number. Thus, 96.4 % of the detected particles are smaller than 0.5 µm, in the range 0.5 - 1 µm are 1.9 %, above 1 µm 1.7 %. Due to the low proportion of particles larger than 0.5 µm, the mass-related fine dust values PM 2.5 and PM 10 are also low. This is also due to the fact that the window side of the classrooms faces a road with hardly any traffic. Even in the case of a five-minute window ventilation phase, only very few particles larger than 0.5 µm are introduced into the room. The influence of the location of the three measuring points is small and does not change the respective characteristic profile of the functions. The sum and density function of the particle size distribution before and after window ventilation is shown in Figure 5 for a characteristic case. The red curve indicates the particle distribution after a five-minute cross-ventilation phase, the black curve shows the distribution before ventilation. The trend shown in the figure is also present in the other measurements. Thus, before ventilation, particle sizes of around 0.3 µm are disproportionately present in terms of number. The peak of the density function of the case shown in Figure 5 is about 0.285 µm. In the range around the peak of 0.255 - 0.316 µm, almost 50 % of all particles of the investigated measuring range are found, whereby the share of particles above 0.5 µm is very low. It is in the single-digit percentage range. Through window ventilation, mainly smaller particles enter the classroom. This shifts the maximum number of particles towards a smaller diameter. Thus, more than 40 % of the particles are in the range of 0.1778 - 0.221 µm. At the same time, however, larger particles, with diameters over 0.4 µm, also enter the classroom through ventilation, which increases their number-related share. When comparing the data of different measurement series, it is noticeable that the proportion of larger particles (> 0.5 µm) during ventilation varies from measurement day to measurement day. At the maximum, up to 10 % of the particles after ventilation were larger than 1 µm.

**Figure 5:**
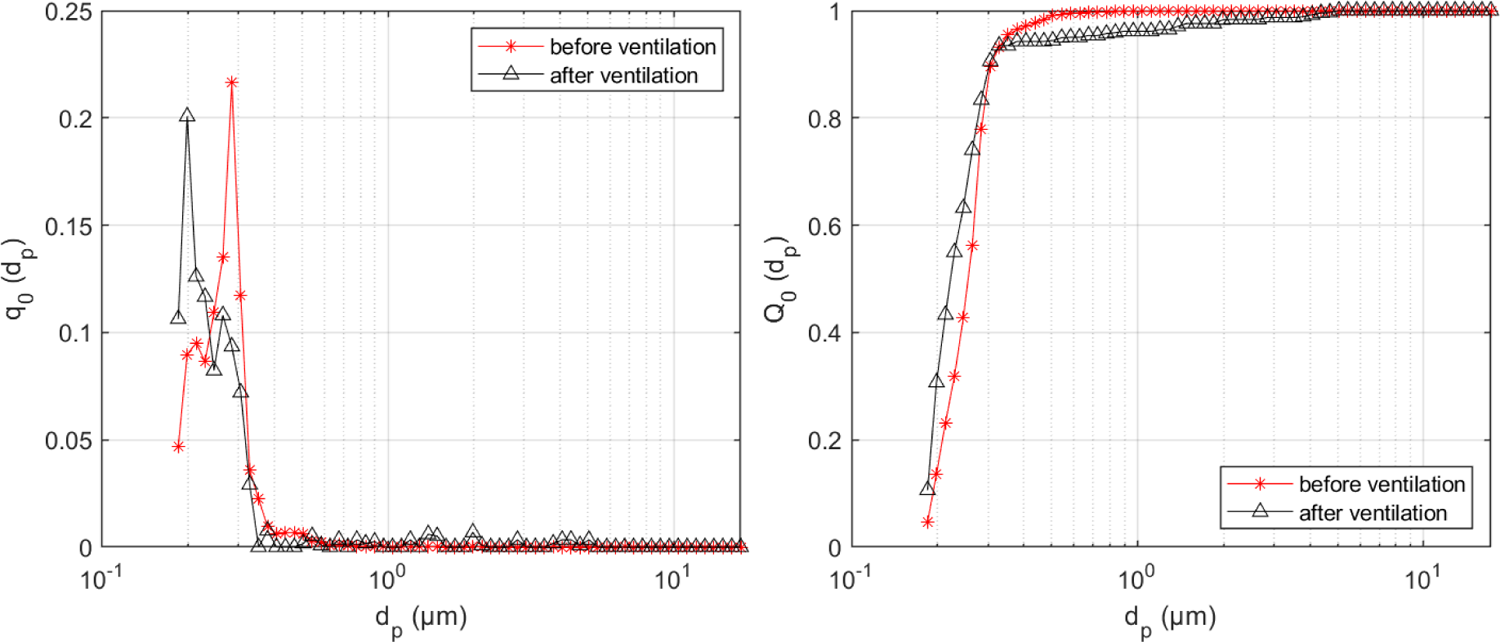
Number-related density *q_0_* and sum distribution *Q_0_* curve of the particle size (*d_p_*) distribution before (red) and after (black) 5 minutes of cross-ventilation

### 3.5 Noise Level

The sound level of the units is a critical factor for classroom use. Table A 2 in the appendix lists the sound pressure levels that the companies have specified for the units. In order to determine the sound levels of the three different units in relation to each other under the same conditions, measurements were taken with a sound analyzer (described in chapter 2.2) at a distance of 2.5 m centrally in front of the unit, with the microphone directed frontally at the unit at a height of 1.08 m. The A-weighted equivalent sound pressure level was measured. The A-weighted equivalent continuous sound level L_Aeq_ was measured over 20 seconds. For the volume flows of 1060 m^3^/h for AP 1 and 1000 m^3^/h for AP 2 and 3 used in this study, values of 36.60 dB(A) for AP 1, 40.45 dB(A) for AP 2, and 38.65 dB(A) for AP 3 were measured. The values with the ranges are summarized in Table 3. The values are mean values from three measurements on two identical units of one type. A survey among the teachers who conducted lessons with the devices over a period of 3 months showed that of the nine teachers, one teacher perceived the noise level as disturbing. The others found it not disturbing (2), rather not disturbing (4) or neutral (2). Thus, the survey is not representative due to the small number of teachers surveyed, but in the opinion of the authors, it is meaningful due to the length of time that the individual teachers surveyed have conducted lessons with the AP. A correlation between the measured noise level and the tuning shows that the perception is strongly subjective. Thus, the teacher who found the level disturbing had AP 3, which was not the loudest according to measurements. In contrast, AP 2, which has a higher level compared to the other two, was not/not at all annoying for the teachers.

**Table 3:** Measured A-weighted equivalent continuous sound level L_Aeq_ of the three tested air purifiers

|  | **Set Volume**<br>**Flow Rate** | **$L_{Aeq}$** | **Range** |
| --- | --- | --- | --- |
| **Air Purifier 1** | 1060 m<sup>3</sup>/h | 36.60 dB(A) | 1.4 dB(A) |
| **Air Purifier 2** | 1000 m<sup>3</sup>/h | 40.45 dB(A) | 0.7 dB(A) |
| **Air Purifier 3** | 1000 m<sup>3</sup>/h | 38.65 dB(A) | 0.6 dB(A) |

In summary, it can be said that the noise level of the units at 1060 (AP 1) or 1000 m^3^/h (AP 2 and 3) does not significantly disturb the teaching process according to the teachers and thus values of up to 40 dB(A) seem unproblematic for APs in schools.

## 4. Sensitivity analysis of the influencing parameters on the particle dose potentially inhaled by individuals

The present study is based on various questions. These include under which conditions the use of large mobile APs is appropriate and which parameters have an influence on the effectiveness of the APs. At the same time, it is of great importance to quantify the effects of individual influencing parameters. A key parameter for evaluating certain measures or influences is the risk of infection. However, determining the risk of infection from airborne aerosols is itself subject to some uncertainty (Lelieveld et al., 2020). For this reason, we do not focus on the risk of infection for our subsequent assessments, but rather on the inhaled particle dose, which represents the number of virus-laden particles ingested by a person. To obtain more precise statements about various influences on the aforementioned inhaled particle dose, it is necessary to vary a large number of parameters under a wide range of conditions. In fact, this would be very time-consuming in a physical experiment. For this reason, we use a calculation model. The following sections address the model assumptions, the test cases studied, the validation, and the results of the parameter study.

### 4.1 Modelling of the inhaled particle dose

The calculation model used in the following is based on the model of (Rietschel & Fitzner, 2008). In the model, an infected person is assumed to emit a constant stream of virus-laden particles into a room. This causes the particles to accumulate in the room and increase the concentration. As a result, the particles accumulate in the room and increase the concentration, while a second healthy person breathes air with the concentration prevailing at the time step. With the help of this model, the concentration of virus-laden particles in a room and the particle dose inhaled by a second person can be mapped as a function of time. The obtained results are shown in the diagrams in Figure 6 for seven different test cases and the boundary conditions specified below.

**Figure 6:**
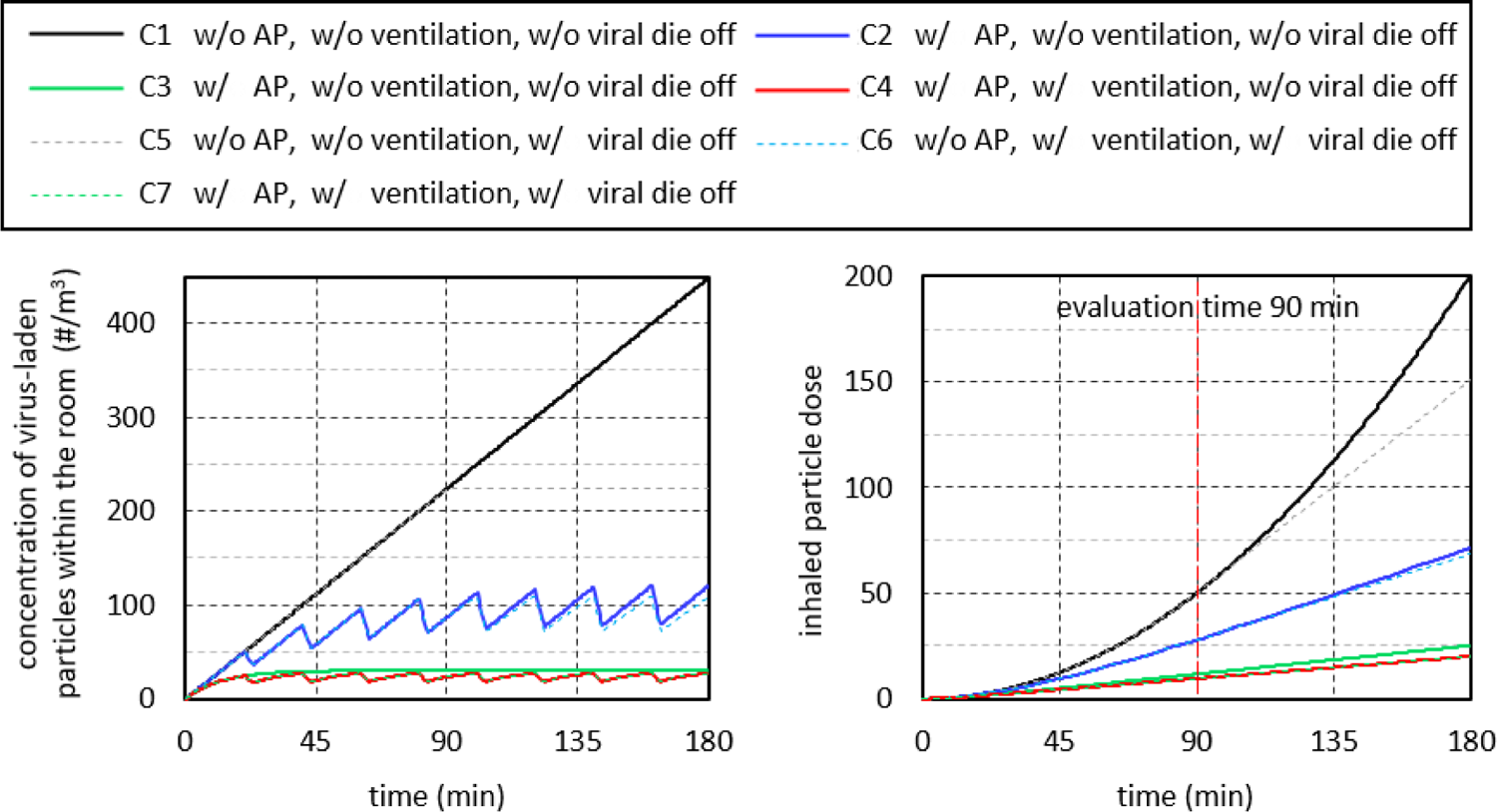
Concentration of virus-laden particles in the room over time (left) and cumulative number of inhaled virus-laden particles (right) for 7 cases. The following conditions are applied: room size 200m^3^, AP volume flow 1000 m^3^/h, ventilation interval 20 min, ventilation duration 3 minutes, ventilation air exchange 50%, viral die-off after 90 min.

The model is based on the assumption that particles are uniformly distributed throughout the room. Consequently, virus-laden particles added to the room air by an infected person with an assumed constant particle flow are uniformly distributed throughout the room. Similarly, if a volume of air in the room is replaced by window ventilation or a certain volume of air is cleaned of particles by an AP, this removal of virus-laden particles leads to a uniform decrease in the particle concentration in the room. The results obtained from this highly simplifying assumption can be considered as a kind of average value. In practice, the particle concentration in a room is location-dependent, which means that the particle dose a person inhales may be larger or smaller than predicted by the model. This may have an influence on the absolute values, but it does not affect the basic conclusions about the effect of different influencing parameters.

### 4.2 Boundary conditions of the test cases

The simplest test case (C1) is the closed room with an air volume of 200 m^3^, in which the particle concentration increases continuously with the virus-laden particles supplied by the breathing air of the infected person (Figure 6 left, black line).

The second test case (C2, blue line) takes into account regular, 20-minute window ventilation for a duration of 3 minutes with a total air exchange of 50 % (ventilation efficiency). These values are based on the empirical data collected during the study on ventilation behavior in schools, although they can be considered an optimistic assumption. When considering ventilation efficiency, the distinction from containment removal effectiveness should be noted. In the curve of the model (blue line), the respiratory particle concentration decreases during the ventilation phases, but not to 50 % of the value that existed before ventilation. There are two reasons for this. First, virus-laden particles continue to be added to the room air by the infected person during ventilation. Secondly, the air exchange in the calculation model takes place proportionally from time step to time step over the duration of ventilation. This means that at a later ventilation time, air is exchanged that has already been diluted with fresh air.

In the third test case (C3), the AP is switched on with a volume flow of 1000 m^3^/h (green line). This removes the number of particles from the room air that are in the filtered air volume. Thus, after a short time, a constant particle concentration is established in the room (left diagram, green line). Here, a state of equilibrium is reached between the number of particles emitted by the infected person and the number of particles removed by the AP.

In rooms without automatic room ventilation, the necessary room air quality, e.g., CO_2_ concentration and humidity cannot be achieved without the window ventilation. Therefore, in practical applications, the operation of an AP must be combined with window ventilation. This results in the fourth test case (C4, red curve). As expected, the concentration of virus-laden particles in the room is slightly below that of the third test case.

For the fifth test case (C5), a virus lifetime of 90 minutes is considered (dashed lines). In a closed room (gray dashed line), the consideration of the virus lifetime has a discernible influence on the inhaled dose of active virus-laden particles when staying in the room for a longer period of time. If regular window ventilation is combined with the consideration of the virus lifetime then case 6 (C6) results (blue dashed line). The last case considered, case 7 (C7), is obtained when the virus lifetime is now taken into account in case four, in addition to the ventilation and the AP (orange dashed line). Here the consideration of the specified virus lifetime has only a marginal influence on the inhaled dose. The reason is that the majority of viruses that die after 90 minutes have already been removed from the room air by ventilation or the AP.

### 4.3 Validation of the calculation model

To validate the computational model, the data measured in classrooms shown previously are used. Hence, the influence of an AP on the particle concentration in a room can be determined from the decay curve of Figure 3. Based on this, the decay curve from the computational model is plotted together with the measured normalized decay curves in Figure 7. For this, an AP volume flow of 1000 m^3^/h, a room size of 186 m^3^, and a constant initial particle concentration were assumed in the model. The curve of the model coincides with the curve of AP 2B and thus lies between the curves of the other cases considered. This shows that the assumptions made in the model are able to represent the influence of the AP on the particle concentration in the room.

**Figure 7:**
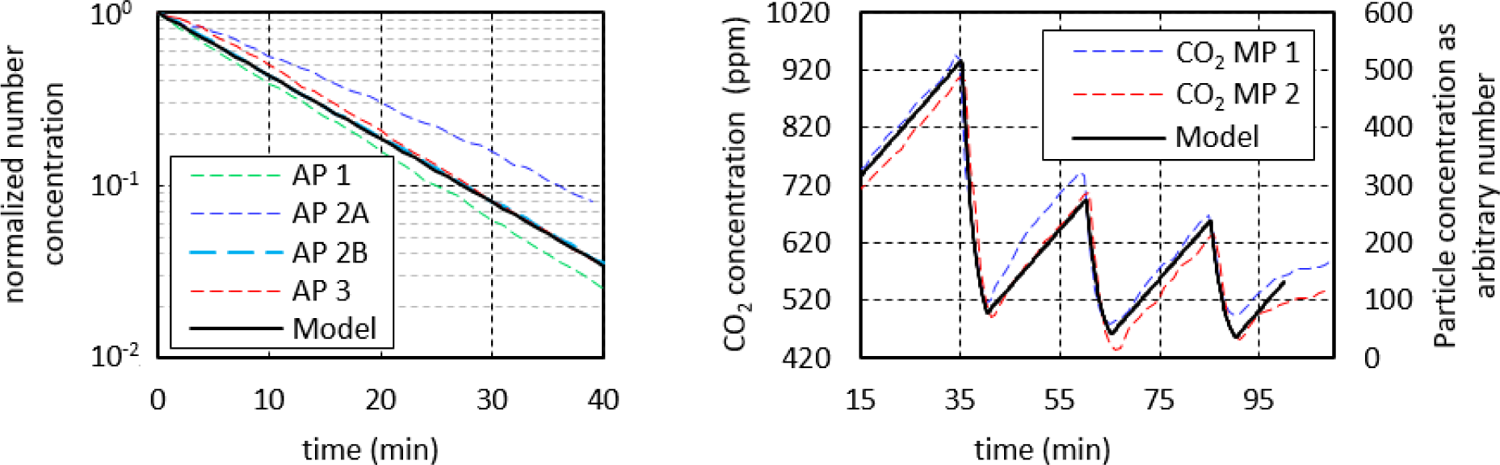
left: Comparison of the AP decay curves between measured and modeled particle concentrations; right: Comparison of the measured CO_2_ concentration with the modeled particle concentration during a lesson.

A second aspect to be considered is the air exchange through window ventilation. During ventilation, a certain proportion of the room air is exchanged with fresh uncontaminated air, thereby removing potentially virus-laden aerosol particles from the room. The change in the CO_2_ concentration in the room can now be used to infer the proportion of air that is exchanged during ventilation (Hartmann & Kriegel, 2020). In this context, a drop in the CO_2_ concentration to the outside air value of 420 ppm represents a complete air exchange. The situation is analogous to the entry of virus-laden particles by an infected person. Here, the increase in the number of virus-laden particles is approximately proportional to the increase in CO_2_ concentration. Thus, the CO_2_ concentration can be used to adjust the calculation model.

The course of the CO_2_ concentration shown in Figure 4 is used to validate the calculation model. For a simple and clear comparison, the values of the particle concentration were normalized to the values of the CO_2_ concentration and reduced by the value 420 ppm. This results in an initial concentration value of 317 ppm. The mean increase in particle concentration between ventilation phases was set to the mean increase in CO_2_ concentration of 0.165 (ppm/s or particles/s). The ventilation efficiency, i.e., the fraction of air exchanged during the ventilation phase, was set to 85 %. The resulting modeled particle concentration is plotted together with the CO_2_ concentration curve in Figure 7 (right).

Here, the significance of the course lies not in the absolute values, which are normalized to the present CO_2_ values. Rather, the significance lies in the gradients, which correctly reflect the change in concentration of the virus-laden particles over the course of the lesson. Rather, the significance lies in the gradients, which correctly reflect the change in particle concentration over the course of the lesson. There are several reasons for the recognizable deviations between measurements and modeling. On the one hand, the increase in CO_2_ concentration, as assumed in the model, is not constant during the lesson; it is influenced, for example, by the intensity of the children’s activity. On the other hand, the CO_2_ concentration in the classroom does not always drop to the same value during the ventilation phases. Despite the controlled ventilation, there are fluctuations in the ventilation efficiency. Furthermore, the measured values are local values in the room and the modeling is an average value.

In fact, it would be easy to increase the agreement between measured CO_2_ course and modeled particle concentration, for example by adjusting the ventilation efficiency individually for the three ventilation phases or by adjusting the increases between the ventilation phases. However, this is not intended at this point. From the results of Figure 7, it can be summarized that the computational model is able to predict real courses of mean particle concentrations in the room air.

### 4.4 Parameter study on the effect of the influencing parameters

Using the previously discussed test cases, the effect of the use of APs on the inhaled particle dose can be determined. At the same time, the effect of influencing parameters on the inhaled particle dose can be determined. In this study, 8 influencing parameters are examined in more detail. These are summarized in Table 4. Considered are the duration of stay, the particle concentration in exhaled air of the infected person, the respiratory flow rate of the infected person as well as of the healthy persons, the virus lifetime, the ventilation interval, the ventilation efficiency, the AP volume flow as well as the room size. In the following effect diagrams, the respective investigated influence parameter is varied in its size. The corresponding variation range is given in Table 4. The variation ranges are based on practical relevance and literature data. In the effect diagrams, the other influencing parameters are set to their standard values as specified in Table 4. For this purpose, values were chosen that either correspond well with the elementary school class scenarios under investigation or that occur with a high probability.

**Table 4:** Overview of the investigated influence parameters with the set standard values and the belonging variation ranges

|  | parameter | unit | set standard | variation range |
| --- | --- | --- | --- | --- |
| P1 | duration of stay | min | 90 | 0 – 180 |
| P2 | particle concentration in exhaled air PCEA | p /cm<sup>3</sup> | 0.1 | 0.1 – 4 |
| P3 | respiratory flow rate | L/min | 5 | 3 – 11 |
| P4 | virus lifetime | min | 90 | 20 – 100 |
| P5 | ventilation interval | min | 20 | 10 – 50 |
| P6 | ventilation efficiency | % | 50 | 10 – 90 |
| P7 | AP volume flow | m<sup>3</sup>/h | 1000 | 400 – 1600 |
| P8 | room size | m<sup>3</sup> | 200 | 50 – 400 |

#### P1 Duration of stay

The duration of stay is one of the parameters that most strongly influence the potentially inhaled dose of virus-laden particles. The right diagram in Figure 6 shows that for case 1 (black line) in an unventilated, closed room, the inhaled particle dose increases exponentially with increasing duration of stay in the room. In this specific case, a doubling of the length of stay leads to a quadrupling of the inhaled particle dose. If regular window ventilation is taken into account in case 2, the value of the inhaled particle dose increases only linearly. Due to the air exchange, the inhaled particle dose is reduced, whereby the advantage over case 1 increases with increasing duration of stay. In case 2, for example, the inhaled particle dose after 45 min is 76 % and after 180 min only 35 % of the inhaled particle dose of case 1. The use of an AP in case 3 (green line) can further significantly reduce the inhaled particle dose. In practice-oriented case 4 (red line), the combination of ventilation and AP, the inhaled particle dose at 180 minutes is reduced to one-tenth compared to case 1.

It should be noted that the inhaled particle dose - and thus also the potential risk of infection - increases continuously in any case with increasing duration of stay in a room. For this reason, the time that several people spend together in rooms, e.g., waiting rooms, should always be kept short. If a common stay over longer periods cannot be avoided, the use of APs significantly reduces the inhaled particle dose, whereby the positive effect strongly increases with increasing duration of stay. In the following considerations of the effects of other influencing parameters, a common duration of stay of 90 minutes is used as a reference value. The background to this is that many schools work with teaching periods of 90 minutes. When interpreting the following results, it must be taken into account that shorter durations of stay would reduce the inhaled particle doses and longer durations of stay would increase the inhaled particle doses. At the same time, the positive effect of using APs would be less pronounced for shorter durations of stay and increasingly more pronounced for longer durations of stay.

#### P2 Particle concentration in exhaled air (PCEA)

The source of airborne virus-laden aerosol particles are infected persons who emit respiratory particles into the environment when breathing or speaking. The concentration or number of particles emitted by a person varies, in some cases greatly, from person to person and depending on the kind of activity. Current knowledge suggests that children emit fewer particles than adults and that quiet breathing emits fewer particles than talking, singing, or shouting (Asadi et al., 2019; Mürbe et al., 2021). Exceptions are so-called “superspreaders”, who are assumed to emit significantly more particles (Asadi et al., 2019). If children breathe calmly, it can be assumed that the PCEA is below the standard value of 0.1 P/cm^3^ assumed here, whereas values above 4 P/cm^3^ can also be reached during singing (Gregson et al., 2021).

In order to determine the effect of the influence of the PCEA on the inhaled particle dose of a healthy person in the room, the PCEA is varied in the range given in Table 4. From the black curve in diagram (1) in Figure 8, it is clear that the particle dose inhaled by a healthy person present in the room increases linearly with PCEA. Thus, due to the wide range of values that the particle concentration can reach, the potentially inhaled particle dose can multiply. Regular ventilation (blue line) and the use of APs (green line) lead to a considerable reduction of the inhaled particle dose, to about 50 % and 25 %, respectively. However, with increasing PCEA, there is also a linear increase in the inhaled particle dose. From these results, it can be concluded that activities that increase PCEA, like singing, should be avoided indoors. In situations where increased PCAE cannot be avoided, ventilation measures and the use of APs are strongly recommended.

**Figure 8:**
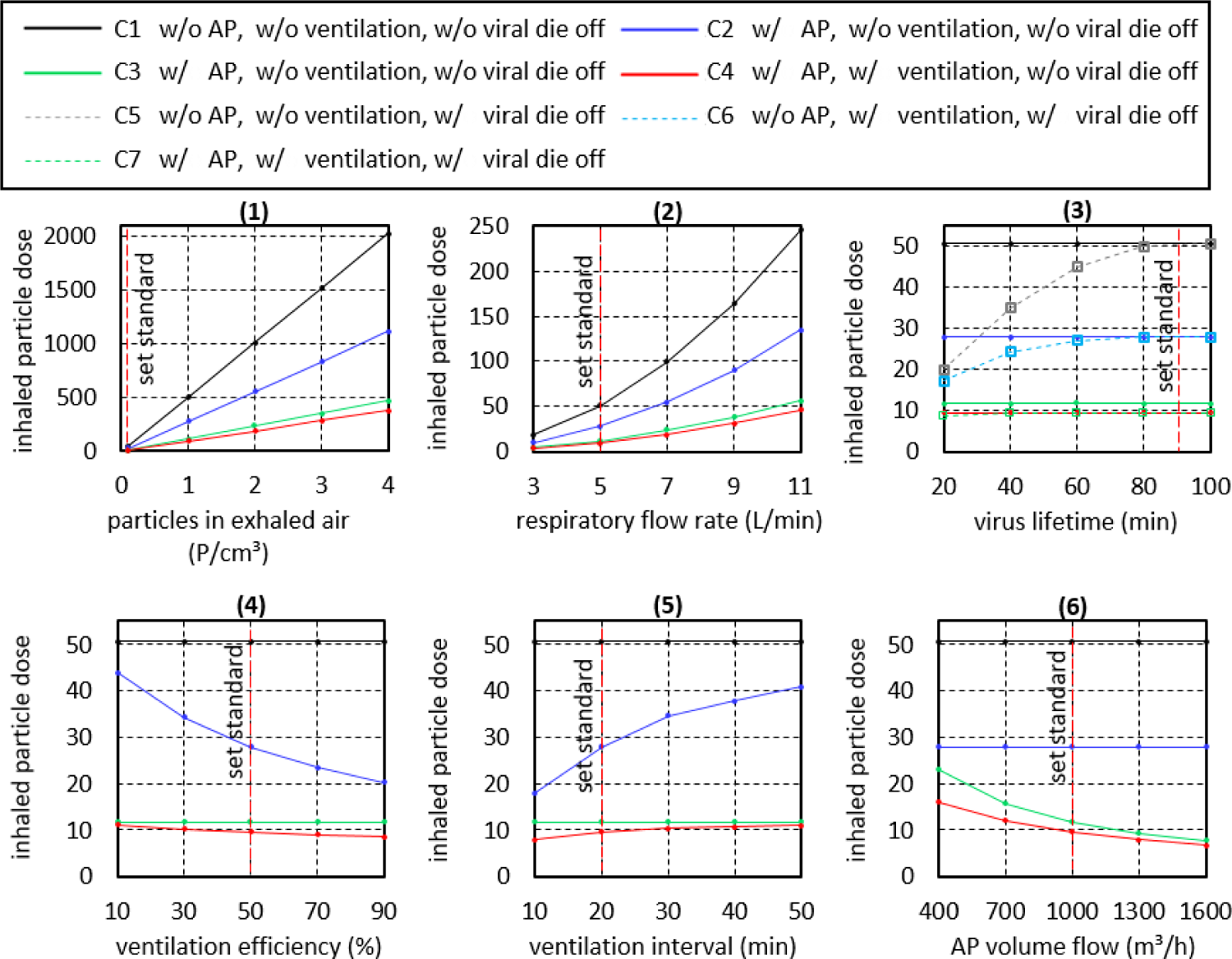
Effect diagrams of the influence parameters of Table 4. Except for the parameter that is varied in each case, the other influence parameters are kept at their set standard value in the individual diagrams. The following conditions are applied: room size 200 m^3^, AP volume flow 1000 m^3^/h, ventilation interval 20 min, ventilation duration 3 minutes, ventilation air exchange 50 %, viral die-off after 90 min.

#### P3 Respiratory flow rate

The respiratory volume flow depends on the age and physical constitution of the persons under consideration. At the same time, the respiratory volume flow is also an indicator of the intensity of the activity, because the respiratory frequency increases with increasing activity. The range of 3 to 11 L/min used here refers to elementary school children, adults reach correspondingly higher values. In the present simulation, the given respiratory volume flow is applied to the infected person and to the inhaling person. Thus, the respiratory volume flow has a “double” influence. Diagram (2) of Figure 8 shows that the inhaled particle dose increases with increasing respiratory volume flow.

If only the respiratory flow rate of the infected person were to be increased, the increase in the particle dose would be linear, just as in the case of the PCEA considered previously. However, since increasingly more air volume is inhaled at the same time, an exponential increase is observed. Although regular ventilation (blue curve) leads to a reduction in the inhaled particle dose, the comparison between closed room, ventilation and AP shows that here only the use of an AP (green line) leads to a substantial reduction in the inhaled particle dose. When considering the aerosol-related risk of infection alone, it can be deduced from the curves that, regardless of the ventilation behavior and the use of APs, sports activities in small rooms should generally be avoided (see also P8 room volume).

#### P4 Virus lifetime

With regard to the risk of infection posed by viruses transported in aerosols, the virus lifetime is an important parameter. However, the lifetime of viruses depends on various boundary conditions, such as air temperature, air humidity or the properties of the emitted aerosol particles. In order to determine the influence of a variation of the viral lifetime on the inhaled particle dose, the lifetime was varied in the range of 20 to 100 minutes (Doremalen et al., 2020). Longer lifetimes were not taken into account because they cannot have any influence at the evaluation time of 90 minutes.

In diagram (3) of Figure 8, the gray dashed curve illustrates that a shortening of the virus lifetime leads to a significant reduction of the inhaled particle dose loaded with living viruses. A very short lifetime of 20 minutes reduces the inhaled particle dose to 20 % of the value without considering virus die-off. At the same time, it must be noted that when regular ventilation (blue line) is used as a comparative scenario, the influence of virus lifetime on the inhaled particle dose is reduced. In the case of simultaneous ventilation and the use of an AP, the virus lifetime has hardly any influence on the inhaled particle dose, because at the time of virus dying, a large proportion of the emitted, virus-laden particles have already been removed from the room air by the AP or by ventilation.

#### P5 Ventilation Interval

The effect of the ventilation interval, namely the distance between the ventilation phases, can be seen from the blue curve in diagram (4) of Figure 8. Smaller intervals between the ventilation phases lead to an increased air exchange during which infectious aerosols are removed from the room. Assuming that 50 % of the room air is exchanged with fresh air during each ventilation cycle (ventilation efficiency of 50 %), the frequently recommended ventilation interval of 20 minutes (IRK, 2020) reduces the inhaled particle dose to 55 % of the value of a closed room. If the ventilation interval is reduced to 10 minutes, the inhaled particle dose is reduced to 35 % of the value of a closed room.

In contrast, the inhaled particle dose approaches the value achieved in a closed room (black line) with increasing distance between the ventilation phases. If we now compare the values of the inhaled particle dose in the case of pure window ventilation (blue curve) with the values achieved with the AP (green curve), the advantage of the AP becomes clear. Even in the case of very short ventilation intervals (assuming the assumed ventilation efficiency), the resulting inhaled particle dose does not fall to the low values achieved by means of AP. In fact, as previously discussed, an AP does not replace the regular performance of window ventilation to improve indoor hygiene. Thus, if one continues to perform regular 20-minute window ventilation in combination with the use of an AP, the inhaled particle dose is reduced even further (red line).

Another important parameter in practice is the ventilation duration. However, since ventilation efficiency is specified in the calculation model, the ventilation duration has only a very small influence on the inhaled particle dose in the calculation model. The effect of the ventilation duration is discussed below in the context of ventilation efficiency.

#### P6 Ventilation Efficiency

The ventilation efficiency, respectively the fraction of room air that is exchanged with fresh air throughout a ventilation phase during window ventilation, is a critical parameter. The ventilation efficiency that can be achieved in practice depends primarily on the prevailing wind speeds and the wind direction. In this case, high wind speeds lead to an acceleration of air exchange, especially during cross-ventilation. Ventilation efficiency also depends on the outdoor temperature, with convective air exchange taking place very quickly, especially at low outdoor temperatures. If the outdoor and indoor temperatures are equal, convective air exchange comes to a standstill. In addition, the room and window geometry as well as the position of the windows play an important role in ventilation efficiency. If the open window area used for ventilation is reduced, this also leads to a reduction in ventilation efficiency. It is advantageous to ventilate through the largest possible open window area. At the same time, the open area should occupy a large part of the room height. Windows with a low height significantly reduce the ventilation efficiency. As a conclusion, it should be noted that in the prevailing poor conditions for air exchange, the duration of ventilation should be increased. Another parameter that has a strong indirect influence on ventilation efficiency is the well-being of the people in the room. It is not only the low temperatures in winter or the drafts that cause discomfort. Disturbing outside noises or pollen in summer can also contribute significantly to a reduction in ventilation behavior. The result is deteriorating air quality in the room. Due to the aforementioned parameters that influence the effectiveness of ventilation, window ventilation can hardly be considered a reliable method for ensuring indoor air quality.

The blue curve in the diagram (5) in Figure 8 illustrates that under the boundary conditions mentioned, a low ventilation efficiency of less than 30 % does not lead to a significant reduction in the potentially inhaled particle dose. In contrast, a high ventilation efficiency of 90 % leads to a reduction of the inhaled particle dose to 40 % of the value in a closed room (black line). It must be noted at this point that, based on the experience gathered in the course of this study, a ventilation efficiency of 90 % is generally not achieved in practical school operation.

Considering the achievable effect of window ventilation and the unreliability of ventilation, the advantage of using an AP becomes clear. An AP reduces the particle concentration in the room in a defined and reliable manner (green line), whereby the potentially inhaled particle dose during AP operation is significantly lower than that during window ventilation. In the practical case, a combination of AP and window ventilation (red line) will be present.

#### P7 Volume flow rate of AP

A frequently discussed parameter is the air exchange rate. In the present case, with constant room volume, the air exchange rate is directly proportional to the AP volume flow. The standard case considered with an AP volume flow of 1000 m^3^/h and a room size of 200 m^3^ corresponds to an air exchange rate of 5 h^-1^. Under these boundary conditions, the use of an AP leads to a decrease in the inhaled particle dose to 20 % of the initial value for the closed room, as diagram (6) in Figure 8 illustrates. An increase of the volume flow (green line) results in a degressive decrease of the inhaled particle dose. Thus, a further increase of the volume flow to 1600 m^3^/h or to an air exchange rate of 8 h^-1^ leads only to a slight further improvement to 15 % of the value of the closed room (black line). It should be noted here that in practice the value of the air exchange rate is limited by the flow velocity that occurs in the room. This is because high flow velocities are perceived as unpleasant. In addition, increasing volume flow rates are associated with increasing noise emissions from the APs; here, a value of 40 dB(A) should not be exceeded.

With decreasing volume flow (green line), the inhaled particle dose approaches the value achieved in closed rooms (black line), whereby even a low volume flow of 400 m^3^/h leads to a halving of the inhaled particle dose. For comparison, the blue line shows the inhaled particle dose in the case of regular ventilation. Under the given boundary conditions, the use of the AP (green line) even with a low air exchange rate of 2 h^-1^ achieves a better value than consequent ventilation (blue line).

#### P8 Room Volume

A parameter with dominating influence on the potential inhaled particle dose is the room volume. The diagram in Figure 9 shows that the inhaled particle dose in closed rooms (black line) increases strongly with decreasing room size. This is plausible because at a constant, continuous particle supply, the particle concentration increases faster in small rooms than in large rooms due to the smaller volume. At the same time, the curve runs towards zero with increasing room size, because in very large rooms - just as outdoors - there is no significant accumulation of virus-laden particles in the given time due to strong dilution of the virus-laden particles with the available air. From this curve, it can be deduced that a risk of infection by aerosols emitted by infected persons exists especially in unventilated, small rooms, as they are predominant in private households. In very large rooms, the risk of infection by airborne virus-laden particles is many times lower due to the strong dilution.

**Figure 9:**
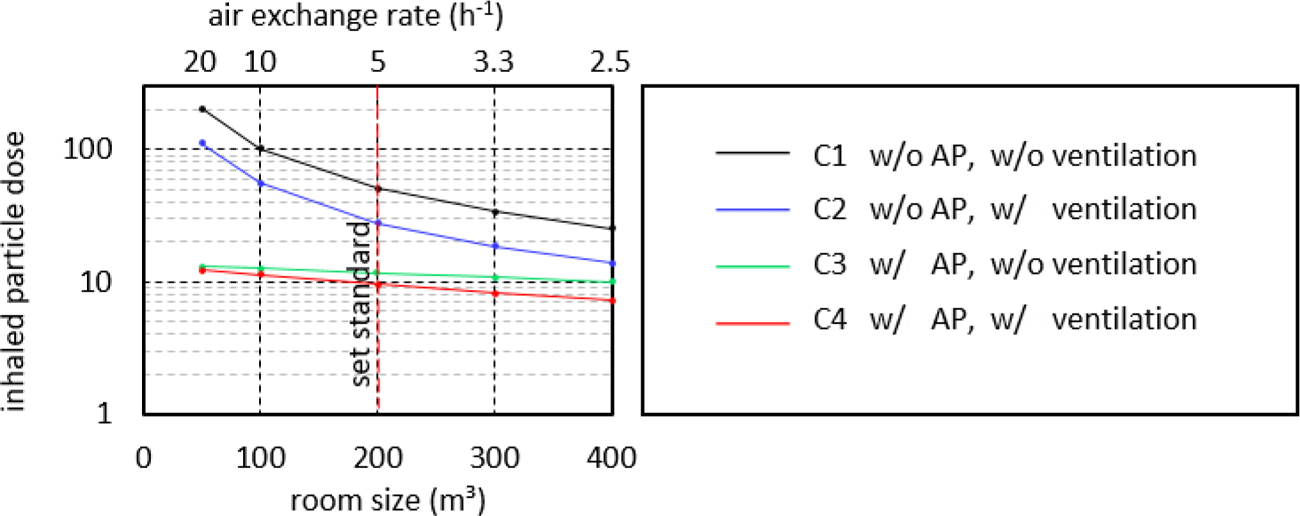
Effect diagram of the room size in logarithmic scale. The other influence parameters are kept at their set standard value in Table 4. The room size is 200 m^3^, the volumetric flow of the AP 1000 m^3^/h.

Regular ventilation (blue line) has a positive effect on the reduction of the inhaled particle dose for all room sizes. Defined window ventilation with the same ventilation parameters leads to an approximately equal proportional reduction of the inhaled particle dose for all room sizes. If one now wishes to reduce the aerosol-related risk of infection in a small room to the low level of large rooms, one must significantly increase the ventilation interval, the ventilation duration, and/or the ventilation efficiency. General guidelines suggest, for example, a ventilation interval of 20 minutes and a ventilation duration of 5 minutes do not guarantee a comparably high air quality for small rooms as can be achieved in larger rooms.

If an AP with a volume flow of 1000 m^3^/h is used (green line), the sum of inhaled particles is significantly reduced compared to window ventilation. Note the logarithmic scaling of the inhaled particle dose. It is interesting to point out that regardless of the room size, the inhaled particle dose remains approximately constant when the AP is used and decreases only very slightly with increasing room size. The reason for the approximately constant curve is that a constant particle concentration is established in the room during operation of an AP over time (cf. Figure 6, left diagram). The value of this particle concentration is independent of the room size. It is reached when the room air filter, due to its volume flow and the prevailing particle concentration, removes exactly as many particles as the infected person emits. Nevertheless, the sum of inhaled particles decreases slightly with increasing room size, since it takes longer for the previously discussed constant particle concentration to set in.

The ratio between the black and green curves in Figure 9 indicates that the positive effect of using an indoor air filter (at the specified airflow rate) is greatest in small rooms. Of course, in this consideration, the air exchange rate is greatest in the small room, and if the same air exchange rate were achieved in the large room, the proportional reduction of the inhaled particle dose would be similar in the large room. It can be concluded, however, that especially in situations where people are in small rooms, the positive effect of the use of APs on the potential inhaled particle dose is particularly significant.

If the curves are placed in the context of the risk of infection caused by aerosols, it is concluded that general statements and recommendations for air exchange rates of APs in rooms are not suitable here. This is because if the inhaled particle dose in a 50 m^3^ and a 500 m^3^ room is to be reduced to approximately the same value, the smaller room requires an air exchange rate of 20 h^-1^, while the larger room only requires an air exchange rate of 2.5 h^-1^ (in the case under consideration). From the curves, it can be deduced that the use of APs is recommended for all room sizes in order to reduce the aerosol-related risk of infection. In small rooms, in general, it should be aimed for high air exchange rates (> 5 h^-1^). In large rooms, APs are also useful, even if the air exchange rates achieved are below 5 h^-1^. If conditions similar to those in the present calculation model are present in practice, an air exchange rate of 2 h^-1^ already halves the particle dose which is inhaled within the first 90 minutes.

## 5 Computational fluid dynamics calculations

In order to assess the flow field inside the classrooms and the effect of APs on this flow and the behavior of aerosols, the Reynolds-Averaged Navier-Stokes (RANS) (Ferziger et al., 2020) equations are solved for a simplified classroom geometry, using computational fluid dynamics (CFD). Temperature variations are accounted for by solving an energy equation and transport of aerosols is modeled as a passive scalar, where the number concentration is considered as a scalar and is evolved using a transport equation.

The flow field inside the classroom is assumed to be incompressible and turbulent and is solved using RANS with the *k* − *ε* closure model. The difference in density due to temperature is modeled using the Boussinesq approximation, which is generally used for ventilation cases. Since the experiments conducted in this study suggest that majority of the aerosol particles have a size of less than 1 µm, the distribution of the aerosols at the scale of a classroom can be modeled using the Eulerian approach, by solving the transport equation accounting for convection-diffusion of the aerosol (Z. Zhang et al., 2021). The effective diffusive constant for the aerosol is the sum of the molecular and turbulent contributions, *D_eff_ = v/Pr + v_t_/Pr_t_*, where Pr = 0.7 and Pr_t_ = 0.85, are the laminar and turbulent Prandtl numbers, respectively, with values that are generally used for the modeling of aerosol transport in ventilation cases (Burgmann & Janoske, 2021). Assuming that the major flow inside a classroom is dominated by the APs which reaches a steady flow over time, a so-called “frozen flowfield” technique is applied to simulate the transient concentration of the aerosol. In this technique, the flow is first calculated assuming a steady-state flow, and then the transient calculation of aerosol transport is performed. This method allows to cut down long simulation time that would otherwise have been taken by a fully transient coupled simulation of the flow field and aerosols. The method described above has been extensively validated with experiments in this study.

### 5.1 Computational domain and case setup

The geometry of the classroom and the setting of tables and purifiers have been selected following the geometry of the experiments (see Figure 2). Simulations of an empty classroom with the AP, student desks, and chairs are done for the validation case. Another set of simulations with 22 students and a teacher has been carried out and validated with experiments by comparing the evolution of CO_2_ and aerosol concentrations. In all the cases, only air circulation by APs is considered and window ventilation is avoided since the flow through a window is subjected to the external local environment which is, as previously discussed, associated with large uncertainties.

The ejection of purified air, its angle with the horizontal plane, and the volumetric flow rate have kept consistent with the specifications of the APs. The volumetric flow of 1060 m³/h is enforced for the case AP1, divided into three jets directed towards the front and top with an angle of 15 degrees to the horizontal plane. For case AP 2 the majority of the flow is towards the side with an angle of 25 degrees with the horizontal and 10 % of the flow is from the front face directed towards the side, with a total volumetric flow rate of 1000 m³/h. For case AP 3, the flow is nearly horizontal and equally divided in front and sides of the air purifier with a volumetric flow rate of 1000 m³/h. The turbulence intensity and the turbulence length scale at the outlet of the air purifier are assumed to be 2.5 % and 0.1 m, respectively. It is assumed that the concentration of aerosols in the air ejected out of the purifier is zero. As the purifier does not remove CO_2_, that has been modeled by mapping the CO_2_ concentration from the purifier inlet to its outlet. Each pupil has been modeled by a volumetric flow rate of 3 m³/h with a concentration of 3 % CO_2_ and a temperature of 307 K. The teacher has been given a slightly higher volumetric flow than students of 3.5 m³/h and 4 % of CO_2_ concentration. The room temperature is assumed to be 298 K. An initial homogeneous concentration of aerosol is assumed throughout the domain and the effect of APs is observed at the points shown in the experiments.

### 5.2 Results of Computational Fluid Dynamics

Firstly, a mesh-independency study is carried out. A hexahedral dominant finite volumes mesh is generated with different mesh resolutions to perform the grid refinement study. The mesh independence criteria and the validation of the model are assessed by comparing the aerosol concentrations at the probe locations mentioned in the experiment. Grid cells with a resolution of 100 mm, 75 mm, and 60 mm as the base size have been used for refinement study, with a total number of cells varying approximately from 2 M, 4.5 M, and 6 M for empty classroom cases and 4.5 M, 9 M and 12 M for cases with students and teacher. The calculation of aerosol and CO_2_ is done by first setting up the flow for the 1800 seconds, followed by transient calculation of aerosols and CO_2_ for another 1800 seconds. The comparison of the evolution of aerosol concentration in an empty classroom at two probes location, one at the mid-point and the other at the far end for different mesh resolutions were in very good agreement with the experimental results.

Figure 10 shows the normalized aerosol concentration plots for experimental and simulation with coarse mesh results. The simulation and experimental plots for cases AP 2B and AP 3 are in good agreement, whereas for cases AP 1 and AP 2A, there are some differences between simulation and experimental results. This difference in results can be caused by several factors, such as the influence of the localized aerosol concentration that might have formed in a real case scenario. It should also be noted that in experimental case AP 2A, the inlet of the purifier is close (0.3 m) to furniture (see Figure A 1 b.) which can have a significant effect on the flow, however, this has not been taken into account in simulations. The *AP 2A exp\** curve in Figure 10 shows the decay curve for the case without furniture near the AP. The decay curve thus approaches the simulated case (blue line). However, there is still a comparatively large difference between the experimental and the simulation results. More detailed findings could be obtained in further investigation through more in-depth experimental flow investigations, also with regard to the influence of furniture and ceiling-mounted objects such as lamps. The effect of furniture was also observed by (Küpper et al., 2019), who showed that obstructing the flow of purifier in an office environment considerably decreased its performance. The result of simulations is, however, in good agreement with the experimental results where the best possible outcome is achieved when the flow is directed towards the front of the room. For case AP 2, in simulation a similar performance is achieved irrespective of its position, however when placed at the center of the long side of the room velocity of air higher than 0.25 m/s was observed near the legs of the students’ desk, which might cause discomfort (Ho, 2021). Further analysis of CO_2_ concentration for the classroom filled with students also showed good agreement with experimental results. As the differences among the results of different mesh resolutions are small, the coarse mesh is used for further simulations.

**Figure 10:**
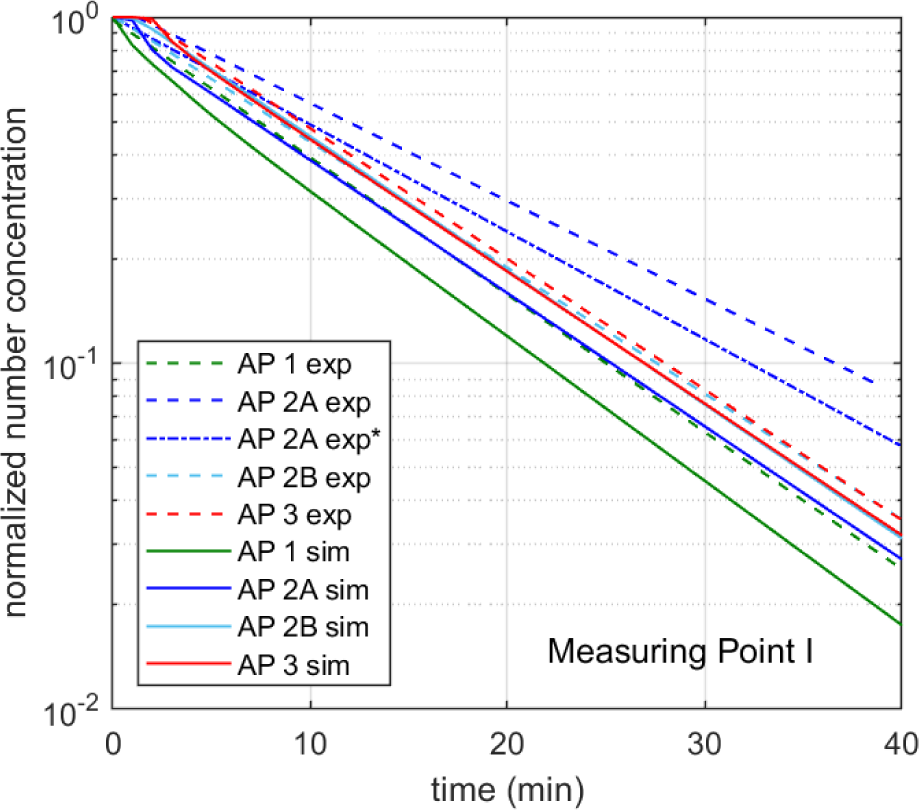
The decay of the normalized aerosol number concentration as a function of time, for the various AP cases, comparison of simulation (sim) with experiment (exp) at measuring point I (see Figure 2). AP 2A exp* shows the decay curve without furniture near the unit.

The evolution of aerosols emitted from the infected person is tracked for all the cases, with the infected person being placed at different locations in the classroom. One case with two students being infected at the first row of the classroom has been simulated to see how the general circulation pattern developed by the purifiers affects the aerosol circulation. The flow pattern develops by AP 1, having front dominant flow is shown in Figure 11. Here, the flow pattern is shown where the air is ejected from the purifier at an angle to the horizontal plane which then has downward circulation. The two students in the front row infected can be observed by the high localized concentration of aerosol. The aerosol concentration after the 30 minutes has been shown where the infected students eject an aerosol concentration of 0.1 #/cm³ particles per second. The spatial concentration of the aerosols can be studied using numerical simulations, which were assumed to be uniform in the mathematical pattern. It was observed that in the AP 1 case the spread was fairly constricted compared to the purifier with dominant side flow which failed to constrict the spread of the aerosol in this extreme case. In the case of AP 3, having equal side and front flow showed similar results to AP 1, which did constrict the spread of the aerosol but was slower compared to AP 1. Generally, the simulation results are in good agreement with the experimental results, as using the AP with front ejection in a classroom shows preferable results compared to the AP with a side dominant flow.

**Figure 11:**
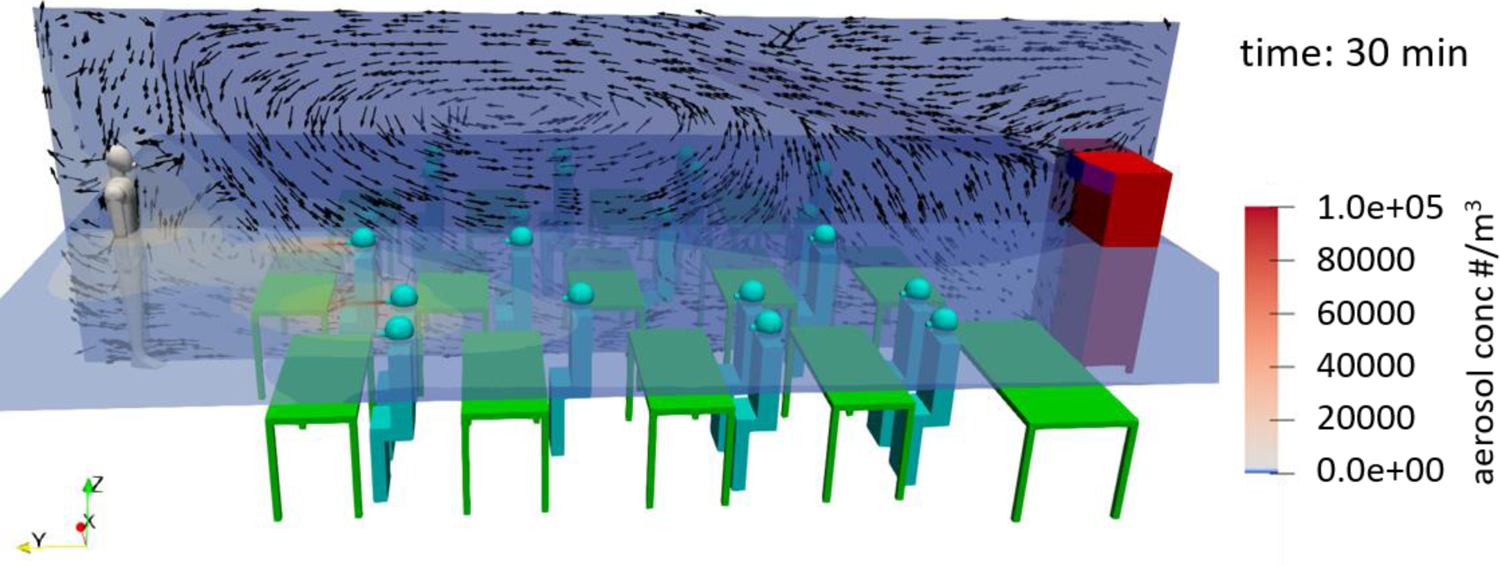
The aerosol concentration after 30 minutes with AP 1. The colors denote the aerosol concentration, whereas the arrows show the local air velocity.

## 6. Summary and Conclusions

The effect of large mobile room air purifiers (AP) on aerosol particle reduction in classrooms (186 m^3^) was investigated in school operation and under reproducible conditions with an aerosol generator. In order to derive a more general conclusion and due to the lack of legal certificates three different high-quality branded APs were tested at a volume flow of around 1000 m^3^/h. The devices differ in the air outlets of the filtered air, which is directed exclusively to the front (AP 1), mainly to the sides (AP 2), and equally to the front and sides (AP 3). In all cases, the air outlet is at a height of more than two meters or at an angle directed upwards. This is necessary to effectively mix the entire classroom with the filtered air and to achieve a uniform dilution of the particle concentration. It can be stated that in a rectangular room, an installation location centrally on the short side of the room opposite of the blackboard, has proven to be advantageous for AP 1 and AP 3, thus half-lives of less than 8 minutes are achieved. AP 2, with the side air outlet, is better placed on the long side of the room. To remove 90 % of the particles from the room, less than 26 minutes (AP 1 and 3) were needed, regardless of the position of the measuring point. Consequently, a large floor-standing unit can demonstrably reduce the concentration of potentially virus-laden particles, measured here in the range 0.178 - 17.78 µm, along the entire length of the room and thus contribute to reducing the risk of infection through airborne viruses indoors.

A critical factor for the suitability and acceptance of such units for use in classrooms is the noise level. For the devices tested, this is not bigger than 40 dB(A) at the volume flow of about 1000 m^3^/h used. A survey among the teachers also showed that this volume and the accompanying air flow had no negative influence on the teaching process. At this point, it must be emphasized once again that high-quality branded devices have been tested here. Since there is no certification of APs so far, the positive results can not necessarily be transferred to other devices. Often, low-cost devices have air bypasses, a very high noise level, and partly also a lack of CE certification. Thus, the selection of a suitable device should not be made without professional advice. In addition, the location of the unit and the furniture in the direct vicinity of the unit have an influence on the performance.

Based on a parameter study conducted using a calculation model it was found that the influencing parameters of room size, duration of stay, and activity of the people in the room have a major influence on the potentially inhaled dose of virus-laden particles. As a result, the use of AP’s has a particularly positive effect in reducing virus-laden aerosols when people are in small rooms for long periods of time and maybe talking, singing, or shouting. Window ventilation reduces respiratory particle concentration and improves air quality in terms of humidity and CO_2_. However, as mentioned above, window ventilation is not a reliable measure to reduce the concentration of virus-laden particles to a defined degree. Compared to conventional window ventilation, the use of well-designed APs achieves a significantly greater reduction in potentially virus-laden particles indoors and does so reliably.

Finally, we have shown the application of computational fluid dynamics (CFD) tools to study the behavior of aerosols in classrooms, which can optimize the placement and operation of APs, as well as study the effect of one or more aerosol emitting people in the room. The results of the CFD calculations are in good agreement with the experiments.

## Data Availability

The authors confirm that the data supporting the findings of this study are available within the article.

## Acknowledgement

Funding of parts of this work by the federal state of Saxony-Anhalt (Germany) is gratefully acknowledged. The authors thank Prof. H.J. Heinze for his help with initiating this study. The authors would also like to thank the head teacher Mrs. Tietge, the caretaker Mr. Rose, the teachers, and the students, without whom this study would not have been possible.

## Conflict of interest

The air purifiers used in the study in the primary school were purchased with funds from the state of Saxony-Anhalt (Germany); there is no participation in the study by the manufacturers. Thus, there is no conflict of interest in this independent study.

## Appendix

**Figure A 1:**
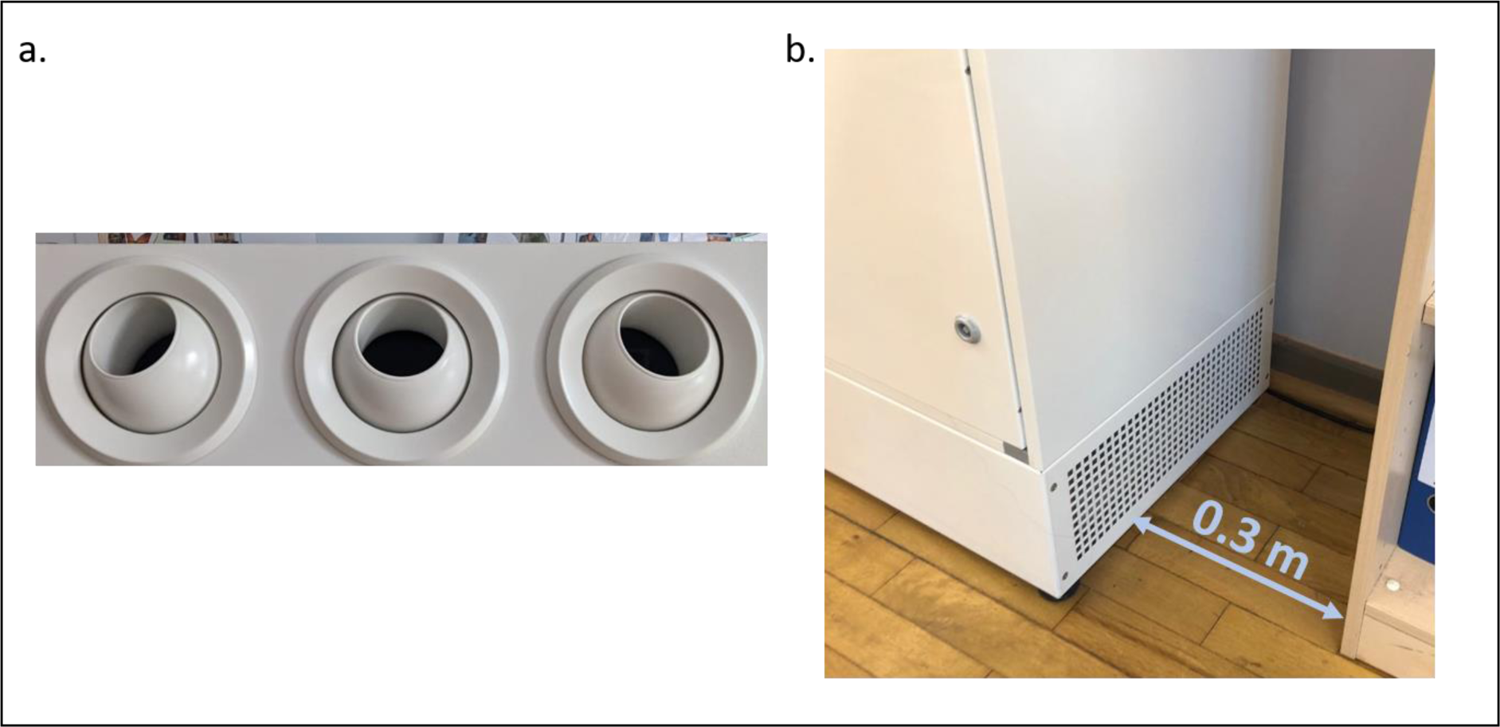
a. Air outlet nozzles of AP 1; b. Side air inlet of AP 2 with the furniture in the classroom

**Table A 1:** Characteristics of the AQ Guard from PALAS according to the manufacturer’s specifications

| Parameters | Values |
| --- | --- |
| Measuring range (size) | 0.178 – 17.78 $\mu\text{m}$ |
| Size channels | 64 |
| Measuring range (number $C_n$) | 0 – 20,000 Particle/$\text{cm}^3$ |
| Measuring range (mass) | 0 – 20,000 $\mu\text{g}/\text{m}^3$ |
| Measuring range air temperature | -20 – +50°C |
| Measuring range air pressure | 700 – 1,100 hPa |
| Measuring range $\text{CO}_2$ [NDIR sensor] | 0 – 5,000 ppm |
| Measuring range for the totality of all volatile organic substances (TVOC) [MOX sensor] | 0 – 60,000 ppb |
| Sampling flow rate | 1 L/min (0.06 $\text{m}^3/\text{h}$) |
| Response time of PM data | 1 s |
| Linearity | 0.95 – 1.05 |

**Table A 2:**
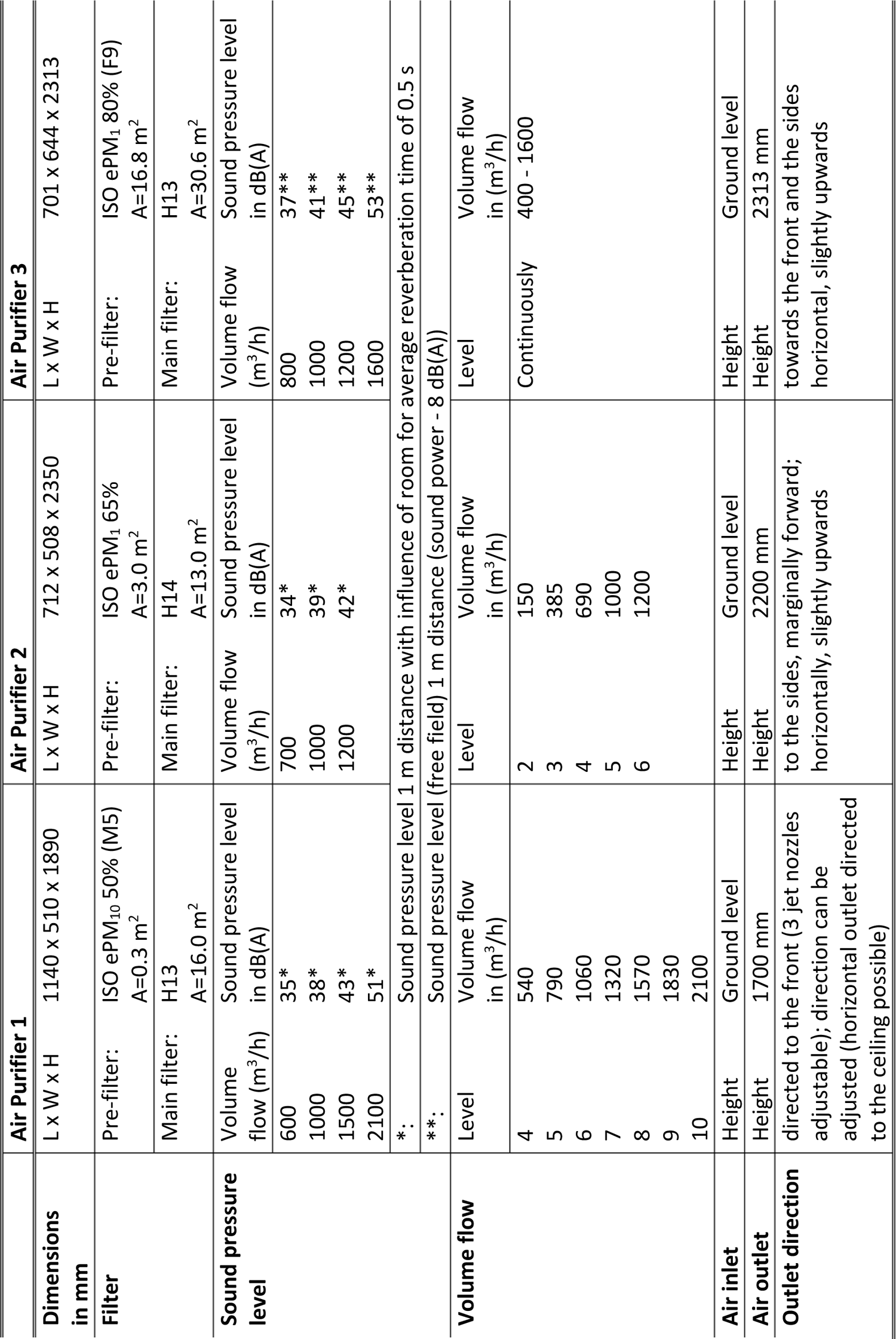
Manufacturer’s data of the three units used in this study

## Notes

### Competing Interest Statement

The authors have declared no competing interest.

### Funding Statement

Funding of parts of this work by the federal state of Saxony-Anhalt (Germany) is gratefully acknowledged. The devices in the study in the school were purchased with funds from the state of Saxony-Anhalt (Germany); there is no participation in the study by the manufacturers.

## References

1. Asadi, S., Wexler, A. S., Cappa, C. D., Barreda, S., Bouvier, N. M., & Ristenpart, W. D. (2019). Aerosol emission and superemission during human speech increase with voice loudness. Scientific Reports, 9(1), 1–10. https://doi.org/10.1038/s41598-019-38808-z

2. ASR A3.7. (2018). Technische Regeln für Arbeitsstätten: Lärm. Retrieved from www.baua.de

3. Bae, S., Kim, H., Jung, T. Y., Lim, J. A., Jo, D. H., Kang, G. S., … Kwon, H. J. (2020). Epidemiological Characteristics of COVID-19 Outbreak at Fitness Centers in Cheonan, Korea. Journal of Korean Medical Science, 35(31), 1–9. https://doi.org/10.3346/jkms.2020.35.e288

4. Brlek, A., Vidovič, S., Vuzem, S., Turk, K., & Simonović, Z. (2020). Possible indirect transmission of COVID-19 at a squash court, Slovenia, March 2020: Case report. Epidemiology and Infection, (March 2020), 2020–2022. https://doi.org/10.1017/S0950268820001326

5. Burgmann, S., & Janoske, U. (2021). Transmission and reduction of aerosols in classrooms using air purifier systems. Physics of Fluids, 33(3). https://doi.org/10.1063/5.0044046

6. Cai, J., Sun, W., Huang, J., Gamber, M., Wu, J., & He, G. (2020). Indirect Virus Transmission in Cluster of COVID-19 Cases, Wenzhou, China. Emerging Infectious Diseases, 26(6), 1343–1345. https://doi.org/10.3201/eid2606.200412

7. Charlotte, N. (2020). High Rate of SARS-CoV-2 Transmission Due to Choir Practice in France at the Beginning of the COVID-19 Pandemic. Journal of Voice. https://doi.org/10.1016/j.jvoice.2020.11.029

8. Curtius, J., Granzin, M., & Schrod, J. (2020). Testing mobile air purifiers in a school classroom: Reducing the airborne transmission risk for SARS-CoV-2. *MedRxiv*. https://doi.org/10.1101/2020.10.02.20205633

9. van Doremalen, N., Bushmaker, T., & Dylan Morris. (2020). Aerosol and Surface Stability of SARS-CoV-2 as Compared with SARS-CoV-1. New England Journal of Medicine, 0–2. https://doi.org/10.1056/NEJMc2004973

10. Fairchild, C. I., & Stampfer, J. F. (1987). Particle Concentration in Exhaled Breath. American Industrial Hygiene Association Journal, 48(11), 948–949. https://doi.org/10.1080/15298668791385868

11. Fears, A. C., Garry, R. F., Roy, C. J., Reed, D. S., Klimstra, W. B., Duprex, P., … Johnson, R. (2020). Comparative dynamic aerosol efficiencies of three emergent coronaviruses and the unusual persistence of SARS-CoV-2 in aerosol suspensions. MedRxiv: The Preprint Server for Health Sciences, 2. https://doi.org/10.1101/2020.04.13.20063784

12. Ferziger, J. H., Peric, M., & Street, R. L. (2020). Computational methods for fluid mechanics (Vol. 4). Springer International Publishing. https://doi.org/10.1007/978-3-319-99693-6

13. GAeF. (2020). Position paper of the Gesellschaft für Aerosolforschung on understanding the role of aerosol particles in SARS-CoV-2 infection, (February). https://doi.org/10.5281/ZENODO.4350494

14. GAeF. (2021). Offener Brief: Ansteckungsgefahren aus Aerosolwissenschaftlicher Perspektive. Retrieved from http://docs.dpaq.de/17532-offener_brief_aerosolwissenschaftler.pdf

15. Gregson, F. K. A., Watson, N. A., Orton, C. M., Haddrell, A. E., McCarthy, L. P., Finnie, T. J. R., … Reid, J. P. (2021). Comparing aerosol concentrations and particle size distributions generated by singing, speaking and breathing. Aerosol Science and Technology, 55(6), 681–691. https://doi.org/10.1080/02786826.2021.1883544

16. Groves, L. M., Usagawa, L., Elm, J., Low, E., Manuzak, A., Quint, J., … Kemble, S. K. (2021). Community Transmission of SARS-CoV-2 at Three Fitness Facilities — Hawaii, June–July 2020. MMWR Surveillance Summaries, 70(9), 316–320. https://doi.org/10.15585/mmwr.mm7009e1

17. Günther, T., Czech-Sioli, M., Indenbirken, D., Robitaille, A., Tenhaken, P., Exner, M., … Brinkmann, M. M. (2020). SARS-CoV-2 outbreak investigation in a German meat processing plant. EMBO Molecular Medicine, 12(12), 1–10. https://doi.org/10.15252/emmm.202013296

18. Hamner, L., Dubbel, P., Capron, I., Ross, A., Jordan, A., Lee, J., … Ball, A. (2020). High SARS-CoV-2 attack rate following exposure at a choir practice. Morbidity and Mortality Weekly Report, 69(19), 606– 610. Retrieved from https://www.cdc.gov/mmwr/volumes/69/wr/mm6919e6.htm

19. Hartmann, A., & Kriegel, M. (2020). Risikobewertung von virenbeladenen Aerosolen anhand der CO 2-Konzentration. *Technische Universität Berlin,* Hermann-Rietschel-Institut, 1–9. Retrieved from http://dx.doi.org/10.14279/depositonce-10361http://dx.doi.org/10.14279/depositonce-10361.3

20. Ho, C. K. (2021). Modeling airborne pathogen transport and transmission risks of SARS-CoV-2. Applied Mathematical Modelling, 95, 297–319. https://doi.org/10.1016/j.apm.2021.02.018

21. HRI/TU Berlin. (2020). Einfluss einer Lüftung auf die Ausbreitung von Partikeln in einem Büro. Retrieved from https://blogs.tu-berlin.de/hri_sars-cov-2/2020/05/12/einfluss-einer-lueftung-auf-die-ausbreitung-von-partikeln-in-einem-buero/

22. IRK. (2020). Einsatz mobiler Luftreiniger als lüftungsunterstützende Maßnahme in Schulen während der SARS-CoV-2 Pandemie Vorbemerkung Lüftungsanlagen und Lüften an Schulen. Retrieved from https://www.umweltbundesamt.de/sites/default/files/medien/2546/dokumente/201116_irk_stellungnahme_luftreiniger.pdf

23. Jang, S., Han, S. H., & Rhee, J. Y. (2020). Cluster of Coronavirus disease associated with fitness dance classes, South Korea. Emerging Infectious Diseases, 26(8), 1917–1920. https://doi.org/10.3201/eid2608.200633

24. Jhun, I., Gaffin, J. M., Coull, B. A., Huffaker, M. F., Petty, C. R., Sheehan, W. J., … Phipatanakul, W. (2017). School Environmental Intervention to Reduce Particulate Pollutant Exposures for Children with Asthma. Journal of Allergy and Clinical Immunology: In Practice, 5(1), 154–159.e3. https://doi.org/10.1016/j.jaip.2016.07.018

25. Katelaris, A. L., Wells, J., Clark, P., Norton, S., Rockett, R., Arnott, A., … Bag, S. K. (2021). Epidemiologic Evidence for Airborne Transmission of SARS-CoV-2 during Church Singing, Australia, 2020. Emerging Infectious Diseases, 27(6), 1677–1680. https://doi.org/10.3201/eid2706.210465

26. Kim, U. J., Lee, S. Y., Lee, J. Y., Lee, A., Kim, S. E., Choi, O. J., … Jang, H. C. (2020). Air and environmental contamination caused by COVID-19 patients: A multi-center study. Journal of Korean Medical Science, 35(37), 1–7. https://doi.org/10.3346/JKMS.2020.35.E332

27. Klompas, M., Baker, M., & Rhee, C. (2020). Airborne Transmission ofSARS-CoV-2: Theoretical Considerations and Available Evidence. Annals of Internal Medicine, 172(11), 766–767. https://doi.org/10.7326/L20-0175

28. Kriegel, M. (2020). Anzahl der mit SARS-CoV-2 beladenen Partikel in der Raumluft und deren eingeatmete Menge, sowie die Bewertung des Infektionsrisikos, sich darüber mit Covid-19 anzustecken. https://doi.org/10.14279/depositonce-10655.4

29. Kriegel, M., Buchholz, U., Gastmeier, P., Bischoff, P., Abdelgawad, I., & Hartmann, A. (2020). Predicted infection risk for aerosol transmission of sars-COV-2. *MedRxiv*. https://doi.org/10.1101/2020.10.08.20209106

30. Kriegel, M., & Hartmann, A. (2020). Ausbreitungsdistanz und-dynamik von Aerosolen in Innenräumen durch Konvektionsströme. Berlin. https://doi.org/10.14279/depositonce-10391

31. Küpper, M., Asbach, C., Schneiderwind, U., Finger, H., Spiegelhoff, D., & Schumacher, S. (2019). Testing of an indoor air cleaner for particulate pollutants under realistic conditions in an office room. Aerosol and Air Quality Research, 19(8), 1655–1665. https://doi.org/10.4209/aaqr.2019.01.0029

32. Laue, M., Kauter, A., Hoffmann, T., Möller, L., Michel, J., & Nitsche, A. (2021). Morphometry of SARS-CoV and SARS-CoV-2 particles in ultrathin plastic sections of infected Vero cell cultures. Scientific Reports, 11(1), 1–11. https://doi.org/10.1038/s41598-021-82852-7

33. Leitliniengruppe Deutschland. (2021). S3-Leitlinie: Maßnahmen zur Prävention und Kontrolle der SARS-CoV-2-Übertragung in Schulen | Lebende Leitlinie: Kurzfassung. AWMF Online, 1. Retrieved from https://www.awmf.org/uploads/tx_szleitlinien/027-076k_Praevention_und_Kontrolle_SARS-CoV-2-Uebertragung_in_Schulen_2021-02_01.pdf

34. Lelieveld, J., Helleis, F., Borrmann, S., Cheng, Y., Drewnick, F., Haug, G., … Pöschl, U. (2020). Aerosol transmission of COVID-19 and infection risk in indoor environments. MedRxiv, 2020.09.22.20199489. Retrieved from https://doi.org/10.1101/2020.09.22.20199489

35. Lendacki, F. R., Teran, R. A., Gretsch, S., Fricchione, M. J., & Kerins, J. L. (2021). COVID-19 Outbreak Among Attendees of an Exercise Facility — Chicago, Illinois, August–September 2020. MMWR Surveillance Summaries, 70(9), 321–325. https://doi.org/10.15585/mmwr.mm7009e2

36. Li, Y., Qian, H., Hang, J., Chen, X., Cheng, P., Ling, H., … Liu, L. (2021). Probable airborne transmission of SARS-CoV-2 in a poorly ventilated restaurant. Building and Environment, 196(January), 14. https://doi.org/10.1016/j.buildenv.2021.107788

37. Lu, Gu, Li, Xu, & Su. (2020). COVID-19 Outbreak associated with air conditioning in restaurant, Guangzhou, China, 2020. Emerging Infectious Diseases, 26(9), 2298. https://doi.org/10.3201/eid2609.201749

38. Ma, J., Qi, X., Chen, H., Li, X., Zhang, Z., Wang, H., … Yao, M. (2020). Exhaled breath is a significant source of SARS-CoV-2 emission. MedRxiv, 1–8. https://doi.org/10.1101/2020.05.31.20115154

39. Makoto, T. (2020). Prediction and Countermeasures for Infection by Virus Contaminated Droplet in Indoor Environment #2. Retrieved June 28, 2021, from https://www.covid19-ai.jp/en-us/organization/riken/articles/article002

40. Mizumoto, K., & Chowell, G. (2020). Transmission potential of the novel coronavirus (COVID-19) onboard the diamond Princess Cruises Ship, 2020. Infectious Disease Modelling, 5, 264–270. https://doi.org/10.1016/j.idm.2020.02.003

41. Moharir, S., Sharath Chandra, S., Goel, A., Thakur, B., Bhalla, G. S., Kumar, D., … Mishra, R. K. (2021). Detection of SARS-CoV-2 in the air from hospitals and closed rooms occupied by COVID-19 patients. MedRxiv, 2020.12.30.20248890. Retrieved from http://medrxiv.org/content/early/2021/01/04/2020.12.30.20248890.abstract

42. Morawska, L., & Milton, D. (2020). It is time to address airborne transmission of COVID-19. Clin Infect Dis, 71(9), 2311–2313. https://doi.org/10.1093/cid/ciaa939.

43. Mürbe, D., Schumann, L., Hartmann, A., Ifrim, L., Von Zadow, D., Seybold, J., … Fleischer, M. (2021). Vergleich der Aerosolpartikelemissionen von Grundschulkindern und Erwachsenen beim Atmen, Sprechen, Singen und Rufen. https://doi.org/10.5281/zenodo.4770776

44. Narayanan, S. R., & Yang, S. (2021). Airborne transmission of virus-laden aerosols inside a music classroom: Effects of portable purifiers and aerosol injection rates. Physics of Fluids, 33(3). https://doi.org/10.1063/5.0042474

45. Papineni, R. S., & Rosenthal, F. S. (1997). The size distribution of droplets in the exhaled breath of healthy human subjects. *Journal of Aerosol Medicine: Deposition*, Clearance, and Effects in the Lung, 10(2), 105–116. https://doi.org/10.1089/jam.1997.10.105

46. Park, J. H., Lee, T. J., Park, M. J., Oh, H. N., & Jo, Y. M. (2020). Effects of air cleaners and school characteristics on classroom concentrations of particulate matter in 34 elementary schools in Korea. Building and Environment, 167(September 2019), 106437. https://doi.org/10.1016/j.buildenv.2019.106437

47. Von Pettenkofer, M. (1858). Besprechung allgemeiner auf die Ventilation bezüglicher Fragen. Retrieved from https://www.luftdicht.de/geschichte/pettenkofer1858.pdf

48. Polidori, A., Fine, P. M., White, V., & Kwon, P. S. (2013). Pilot study of high-performance air filtration for classroom applications. Indoor Air, 23(3), 185–195. https://doi.org/10.1111/ina.12013

49. Probst, W. (2003). Arbeitswissenschaftliche Erkenntnisse - Forschungsergebnisse für die Praxis, Bildschirmarbeit - Lärmminderung in Mehrpersonenbüros. 1. Auflage. Bremerhaven: Wirtschaftsverlag NW Verlag Für Neue Wissenschaft GmbH 2003, 1–27. Retrieved from https://www.baua.de/DE/Angebote/Publikationen/AWE/AWE124.html

50. Qian, H., Miao, T., Liu, L., Zheng, X., Luo, D., & Li, Y. (2021). Indoor transmission of SARS-CoV-2. Indoor Air, 31(3), 639–645. https://doi.org/10.1111/ina.12766

51. Rietschel, H., & Fitzner, K. (2008). *Raumlufttechnik Band 2: Raumluft-und Raumkühltechnik* (16th ed.). Springer-Verlag Berlin Heidelberg. https://doi.org/10.1007/978-3-540-68267-7

52. Shen, Y., Li, C., Dong, H., Wang, Z., Martinez, L., Sun, Z., … Xu, G. (2020). Community Outbreak Investigation of SARS-CoV-2 Transmission among Bus Riders in Eastern China. JAMA Internal Medicine, 180(12), 1665–1671. https://doi.org/10.1001/jamainternmed.2020.5225

53. Stadnytskyi, V., Bax, C. E., Bax, A., & Anfinrud, P. (2020). The airborne lifetime of small speech droplets and their potential importance in SARS-CoV-2 transmission. Proceedings of the National Academy of Sciences of the United States of America, 117(22), 11875–11877. https://doi.org/10.1073/pnas.2006874117

54. UBA. (2021). Lüftung, Lüftungsanlagen und mobile Luftreiniger an Schulen. Retrieved from https://www.umweltbundesamt.de/themen/lueftung-lueftungsanlagen-mobile-luftreiniger-an

55. VDI 6022. Ventilation and indoor-air quality - Hygiene requirements for ventilation and air-conditioning systems and units (VDI Ventilation Code of Practice) (2018). Germany.

56. VDI2081. Air-conditioning - Noise generation and noise reduction (2019). Düsseldorf, Germany. VDMA. (2020). *Raumklima deutscher Schulen erfordert Handlungsbedarf*. Retrieved from https://aig.vdma.org/documents/105643/52060450/PI_Raumklima deutscher Schulen erfordert Handlungsbedarf_1600343834592.pdf/ed94cf1d-161c-524c-cbb7-8a1c9e6e1f83

57. WHO. (2020). Transmission of SARS-CoV-2: implications for infection prevention precautions. Scientific brief, 09 July 2020. Retrieved from https://www.who.int/news-room/commentaries/detail/transmission-of-sars-cov-2-implications-for-infection-prevention-precautions%0Ahttps://bityli.com/j84ms

58. WHO. (2021). Roadmap to improve and ensure good indoor ventilation in the context of COVID-19. Retrieved from https://www.who.int/publications/i/item/9789240021280

59. Zhang, R., Li, Y., Zhang, A. L., Wang, Y., & Molina, M. J. (2020). Identifying airborne transmission as the dominant route for the spread of COVID-19. Proceedings of the National Academy of Sciences of the United States of America, 117(26), 14857–14863. https://doi.org/10.1073/pnas.2009637117

60. Zhang, Z., Han, T., Yoo, K. H., Capecelatro, J., Boehman, A. L., & Maki, K. (2021). Disease transmission through expiratory aerosols on an urban bus. Physics of Fluids, 33(1). https://doi.org/10.1063/5.0037452

